# Clinically Grounded AI-Scribing in Psychotherapy: Benchmarking LLMs Against Expert Documentation in the iCARE Framework

**DOI:** 10.1101/2025.06.25.25330252

**Authors:** Prottay Kumar Adhikary, Suruchi Singh, Sahajpreet Singh, Rashmi Choudhary, Charu Saxena, Prachi Chauhan, Panna Sharma, Salam Michael Singh, Swati Kedia Gupta, Koushik Sinha Deb, Tanmoy Chakraborty

## Abstract

**Background:** AI-scribes based on large language models (LLMs) are being rapidly adopted in healthcare, yet their use in psychotherapy remains limited because most systems produce brief, administratively oriented notes that miss the interpretative and contextual nuance essential to mental-health documentation. We argue that AI documentation for psychotherapy should instead be built on a comprehensive structure of history, clinical evaluation, and risk assessment that can subsequently be condensed into any brief format.

**Objective:** To this end we introduce iCARE (**I**dentifying information, **C**hief concerns and clinical history, **A**ssessment and analysis, **R**isk identification, **E**valuation of progress and action plan), a structured, clinically grounded, 17-section documentation format developed through iterative review by psychiatrists and clinical psychologists.

**Methods:** Using the publicly available HOPE dataset, we created iHOPE, a corpus of 174 re-transcribed, speaker-diarized therapy sessions with expert-written gold-standard notes in the iCARE format. We benchmarked eleven contemporary LLMs (nine open-source and two proprietary) in zero-shot and one-shot settings, evaluating outputs with lexical (BLEU, METEOR, ROUGE-L) and semantic (BERTScore, BLEURT, InfoLM) metrics and with TRACE (Trustworthiness, Relevance, Accuracy, Comprehensiveness, Expression), a five-domain human-evaluation framework we designed for mental-health documentation.

**Results:** Closed-source models consistently outperformed open-source ones, with GPT-4o-mini achieving the best overall automatic-evaluation scores and Gemini Pro excelling at interpretative sections such as crisis-marker identification. All models struggled with temporal reasoning, such as past-session and next-session details, and lexical overlap with human notes was low despite strong semantic alignment. In blinded expert review, clinical preference did not always mirror automatic benchmarks, with Mistral, a smaller open model, emerging as a surprise favourite.

**Conclusions:** The iCARE format, iHOPE dataset, and TRACE framework together provide a foundation for clinically valid, therapist-centred AI documentation that assists rather than replaces clinicians.

## Introduction

### Background

Large Language Model (LLM) based documentation agents, often called “AI-scribes” have found rapid adoption in healthcare, as they reduce the clerical burden and improve efficiency [1]. These agents are often integrated into the clinic architecture and Hospital Information System (HIS), where they automatically record, transcribe and summarize clinical discourse, allowing clinicians to devote more attention to direct patient care [2]. Yet, use of such AI-scribes remains significantly constrained in mental healthcare, as most agents lack the nuanced understanding necessary to summarise a client-therapist discussion [3]. While clinical notes in most medical disciplines focus on objective data (symptoms, duration, lab reports etc.), mental health evaluations remain significantly more subjective and interpretative [4]. In psychology, where narratives and emotions form the core of clients’ suffering, considerable clinical judgement is required to attach weights and biases to the clients’ “talk” to separate out the important and the imminent from the incidental and anecdotal [5].

Developing AI-scribes for therapy note-taking is also complicated by the plethora of psychotherapy note types and formats available [6]. Psychotherapy notes began historically as unstructured personal reflections of the therapists [7], and early systematic approach can be traced back to the work of Adolf Meyer in 1918 [8]. These “Meyerian” case-notes, though structured, were not brief. Rather, Meyer recommended a comprehensive life-course approach, where understanding came from the totality of client’s development, family and social context, habits, personality, precipitating stresses, and current mental status. Structured “micro-note” formats such as SOAP (Subjective, Objective, Assessment, Plan) [9], DAP (Data, Assessment, Plan), BIRP (Behaviour, Intervention, Response, Plan), DART (Description, Assessment, Response, Treatment) etc. [10], were introduced much later in the 1980s, with the advent of multi-disciplinary, hospital based, insurance covered practice. These micro formats, currently used globally, standardise therapy documentation, save therapist time, and facilitate rapid comprehension by other team-members, including general physicians, social workers, nursing staff, and insurance providers. They also, by design, exclude sensitive client information, as these notes may be accessed by a broader group of health-system-associated professionals. The Health Insurance Portability and Accountability Act (HIPAA) specifically makes a distinction between personal “therapy notes” that include such sensitive details and “clinical notes” that are more process/outcome centred [11]. Notes also vary according to the therapy life-cycle, with “intake notes” at initiation, “session notes” after each session and “therapy summary” or “termination notes” at therapy completion.

In practice, most therapists maintain a detailed case-history for their own reference, while also generating a structured summary for administrative, regulatory and insurance purposes. Psychological associations across USA [12], UK [13] and Australia [14], as well as most textbooks on the subject [10,6], accordingly provide extensive guidance on clinical history elicitation as well as on structured brief notes. From the therapist’s perspective, the detailed clinical history (as a part of intake note) still holds unparalleled value, as it helps the clinician construct a “world model” [15] of the client. Clinical history provides the scaffold that gives meaning to the client’s current narrative; and without it, understanding remains incomplete or even erroneous. These clinical history notes, rooted in the Meyerian tradition, are typically extensive and demand the greatest therapist time for reflection and summarization. SOAP and similar brief notes, in comparison, are cognitively much less demanding for the therapist. Using LLMs solely for SOAP/DAP note generation [16], while useful for administrative efficiency, misses the bigger problem of the field and under-utilises the potential of current AI models. Automated scribing needs to be capable of capturing the nuance and contextual complexity of the therapeutic session itself, to have any meaningful clinical value.

We, therefore, argue that AI documentation for psychotherapy must be grounded in a “mega-structure” of comprehensive history, clinical evaluation, and risk assessment – elements that are of critical value to both therapist and client. This mega-structure of client information can be subsequently “quantised” to SOAP/DAP or any other format, according to administrative or clinical needs. While SOAP/DAP notes are not inter-convertible and cannot be combined across multiple sessions to provide “handover notes” (when client is shifted to another therapist) or “therapy termination notes” (summary at the end of therapy), a Meyerian mega-structure can. To this objective, in this paper, we propose, develop, and test a novel psychotherapy documentation system, using currently available LLMs.

### Related Work

The literature on AI-assisted documentation in psychotherapy (summarized in Multimedia Appendix 2) is relatively new, but it shows a clear progression from early algorithmic digital support tools toward increasingly sophisticated automated LLM systems. For example, in an early work from 2015, researchers used a Tele-Board MED software system during CBT sessions and attempted automatic capture of session content with collaborative documentation [17,18]. In contrast, current papers focus on domain-aligned documentation [19] and go beyond the use of conventional summarization models such as BART and T5 [20]. Recent research deploys nuanced LLM-based strategies, such as planning-guided summarization [21], domain-informed filtering [22], and aspect-specific note generation [23] to capture various facets of therapy documentation. The two large-scale studies of 2026 stand out, as they test AI-deployment at scale, for routine psychotherapy. Ward et al. [24] evaluated AI notetaking across 853,605 appointments, 8,724 providers, and 189,530 patients, and reported 25% faster note completion, 8.2 hours earlier note submission, and reduced after-hours work with AI. Similarly, McCrudden et al. [16] conducted a 1-year retrospective evaluation of a commercial virtual mental health platform, where 162 full-time and 1,366 contractual providers generated more than 286,000 clinical notes; and found AI-adoption to be high and stable. Available evidence, therefore, suggests AI-powered documentation to be promising, feasible and acceptable to therapists. While concerns have been raised regarding hallucination, semantic drift, over-compression of nuanced therapeutic material, and insufficient human validation in these papers [25]; commercial deployment of such AI-systems are rapidly underway in most parts of the world. This radical transformation of health systems with AI creates an urgent need for more rigorous research, especially in mental health care, where clinicians bear an ethical responsibility to protect vulnerable clients.

### Study Objectives

In this paper, we evaluate the role of LLMs in automated psychotherapy documentation. Our work, therefore, is built on four core objectives:

- We introduce iCARE (Identifying Information, Chief Concerns and Clinical History, Assessment and Analysis, Risk identification, Evaluation of progress and Action plan), a novel, comprehensive, structured, psychotherapy documentation format for AI-scribing. iCARE is designed to be information rich, clinically relevant and familiar to mental health professionals.
- We develop iHOPE, a corpus of gold-standard psychotherapy notes in iCARE framework, written by a team of mental health experts from the publicly available HOPE dataset [31]. This dataset will serve as a foundation for training LLMs in detailed therapy documentation.
- We introduce TRACE (Trustworthiness, Relevance, Accuracy, Comprehensiveness, and Expression), an evaluation framework for health documentation quality, purposely tailored for mental health care settings.
- We finally evaluate through TRACE the performance of 11 contemporary LLMs in automated psychotherapy session documentation in the iCARE format, using the iHOPE dataset, relative to expert-generated gold-standard session notes.

## Methods

### Overview

The experimental design of this study is organised in five key steps, each contributing towards improving psychotherapy documentation through use of AI. Figure 1 provides an overview of the methodology of the study.

**Figure 1.**
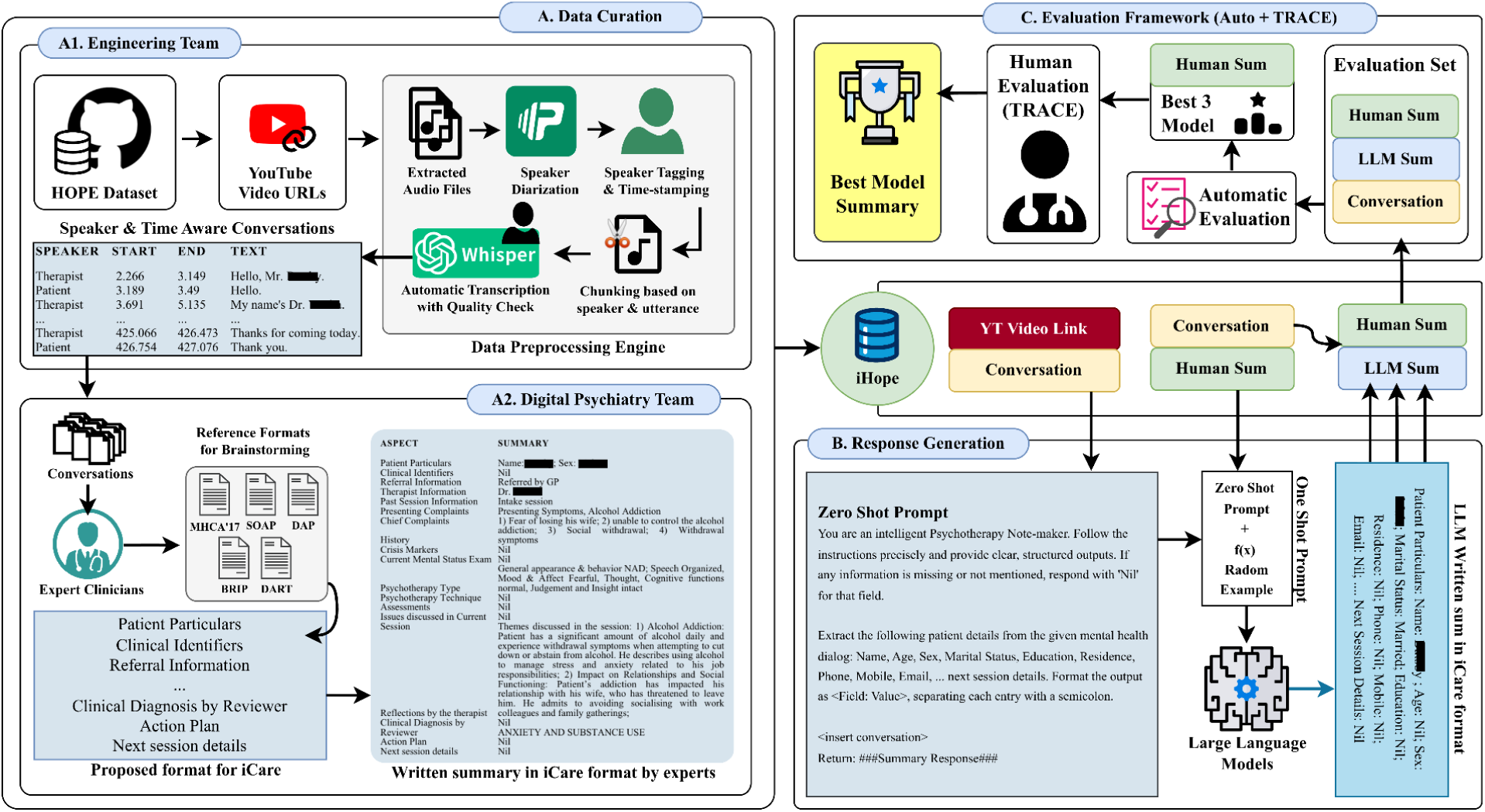
An overview of the methodology employed in this study. (A1) The HOPE transcript generation phase began with data cleaning, speaker tagging and segmenting the full conversations into meaningful chunks of audio files. (A2) followed the proposed iCARE format resulting in iHOPE dataset. (B) Benchmarking leveraged diverse prompting techniques applied to multiple state-of-the-art language models. (C) Evaluation metrics were employed to assess model performance rigorously, incorporating quantitative metrics alongside qualitative assessments to ensure alignment with clinical and therapeutic standards.

### Development of the iCARE Format for AI-Psychotherapy Documentation

The development process for iCARE (Identifying Information, Chief Concerns and Clinical History, Assessment and Analysis, Risk identification, Evaluation of progress and action plan) included an extensive literature review of global psychotherapy documentation practices [6,7,10,32-34], organizational guidelines (e.g. American Psychological Association), laws and acts (e.g. Health Insurance Portability and Accountability Act, USA) [11], as well as various institutional reporting templates [12]. We also reviewed all structured brief-reporting formats, such as POMR, SOAP, DAP, BIRP, etc. [35] to determine commonalities and differences. The initial set of collated components were then independently reviewed by five teaching and clinical faculty members, including psychiatrists and clinical psychologists. The items of the final structured format were iteratively refined by the research team through merging overlapping components and by removing less essential ones, as per agreement between experts.

### Development of the iHOPE Dataset

For this research, we utilised the publicly available HOPE dataset [36], which contains a total of 174 pre-recorded therapy session videos in English language, sourced from YouTube. These sessions represent various phases of psychotherapy (e.g. intake, assessment, follow-up) and cover a range of mental health conditions like depression, anxiety, post-traumatic stress disorder, substance use disorders etc. The HOPE Dataset has been used in various studies [31,37-39] as well as by us, in our previous work [19]. We reprocessed the HOPE dataset to generate complete, high-accuracy transcripts using WhisperX [40] by OpenAI [41], as available transcripts were often found to be incomplete. We additionally performed speaker diarization using pyannote [42] followed by speaker assignment to identify which diarized speaker corresponded to the therapist and which corresponded to the patient. We reviewed each transcript alongside the original video to correct transcription errors and improper speaker tags, filled in missed dialogues, and removed irrelevant/repeated content while preserving the original timestamps to maintain temporal integrity. This rigorous manual validation ensured a high quality of the generated iHOPE transcripts. Using these iHOPE transcripts, an expert panel of three psychologists and two psychiatrists independently reviewed and generated detailed notes using the iCARE framework for all the 174 therapy sessions. The process was supervised by a faculty of psychiatry with 18 years of clinical experience and a faculty of clinical psychology with 14 years of clinical experience who randomly transcribed 10% (n=18) of the sessions. This manually processed corpus of session notes resulted in the human gold standard reference dataset (iHOPE) against which all model outputs were measured.

### Session Note Generation With LLMs

We evaluated a total of 11 LLMs, including both open-source models, Gemma [43], Llama 3.1 [44], Llama 3 [44], MentalLLama [45], Mistral [46], Orca 2 [47], Phi 3 [48], Vicuna [49], and Zephyr [50] and proprietary models, GPT-4o-mini [51] and Gemini Pro [52]. Multimedia Appendix 1, section S1.2 provides a detailed description of the models used. For summary generation with these models, we used default model parameters for closed source models, while for open-source models, the maximum number of new tokens was set to 100. Because each of the 17 iCARE sections was generated independently using a dedicated, section-specific prompt (Multimedia Appendix 1, section S1.1), and the corresponding gold-standard reference for each aspect is short and structured, a 100-token limit was sufficient to fully capture the content of a section while discouraging the open-source models from over-generating or drifting off-topic, and it kept output lengths broadly comparable across models. All open-source models selected were in the 7-8 billion parameter range for a fair and balanced comparison. For prompt design, both teams (mental health professionals and computer science/AI experts) worked in collaboration to create precise prompts that specified task essentials, boundary conditions (what not to include) and desired output formats for each of the 17 sections of the iCARE format. (Multimedia Appendix 1, section S1.1) All LLMs were tested under two experimental conditions. In zero-shot, models were given the iHOPE transcripts and prompted to generate a summary note in the iCARE format. One-shot prompting, in contrast, simulated basic training by providing a single example of human summary of the transcript in the iCARE format.

### Benchmarking Model Performance

We explored lexical and semantic-based methods to systematically evaluate the generated responses. Lexical similarity metrics BLEU [53], METEOR [54], ROUGE-L [55] helped us assess elements like extracted nouns (e.g., patient details). For more nuanced aspects of the summaries (risk assessment, therapist reflections, etc.), we utilised BERTScore [56], BLUERT [57], and InfoLM [58] to capture the semantic meaning of the responses. Unless stated otherwise, all scores are represented as real numbers ranging from 0 to 1, with 1 being the highest possible score. See section S1.3 of Multimedia Appendix 1 for detailed information on automatic evaluation metrics.

### Human Evaluation of LLM Documentation

To determine an appropriate human evaluation framework for psychotherapy documentation, we reviewed several published metrics, including QUEST [59], TN-Eval [60], XLingHealth [61], MEDIC [62], CLEAR [63], and HumanELY [64]. Systematic reviews [59,64-66], however, note substantial heterogeneity in these evaluation criteria with domains of trust and bias being least represented. Additionally, the long-form and reflective nature of psychotherapy notes differ significantly from the needs of most other medical documentation, where these evaluation frameworks have been primarily used. We, therefore, additionally conducted discussion sessions with multidisciplinary experts, from computer science and mental health, and derived at a five-domain evaluation metric: TRACE (Trustworthiness, Relevance, Accuracy, Comprehensiveness, Expression), better suited for evaluation of therapy and psychiatry documentation (Table 1). Participating mental health faculty were requested to “share parameters on which they evaluate the quality of clinical notes written by trainee residents in psychiatry and psychology,” and teaching methods for guiding “good” session notes, which formed the basis of the finally selected domains. All five domains of the metric are rated on a Likert scale from 1 to 5, where 1 is the lowest and 5 is the highest possible quality.

**Table 1.**
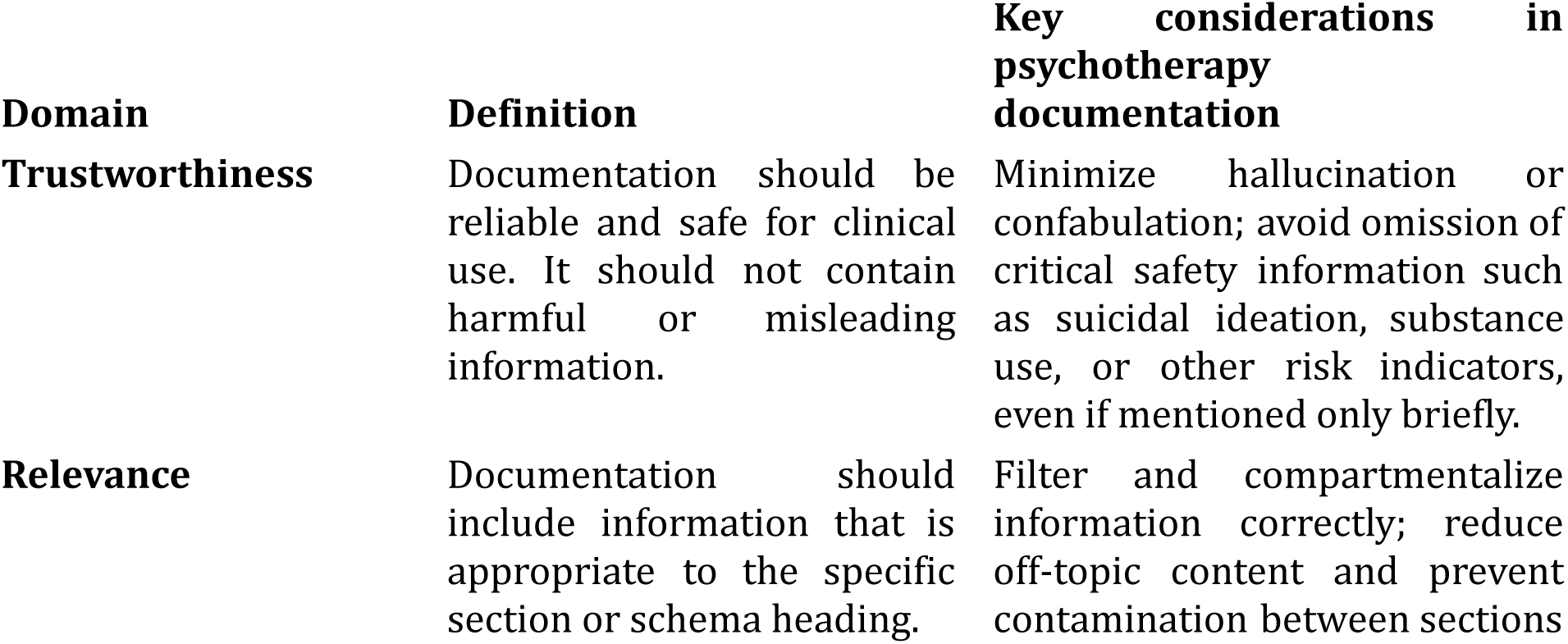
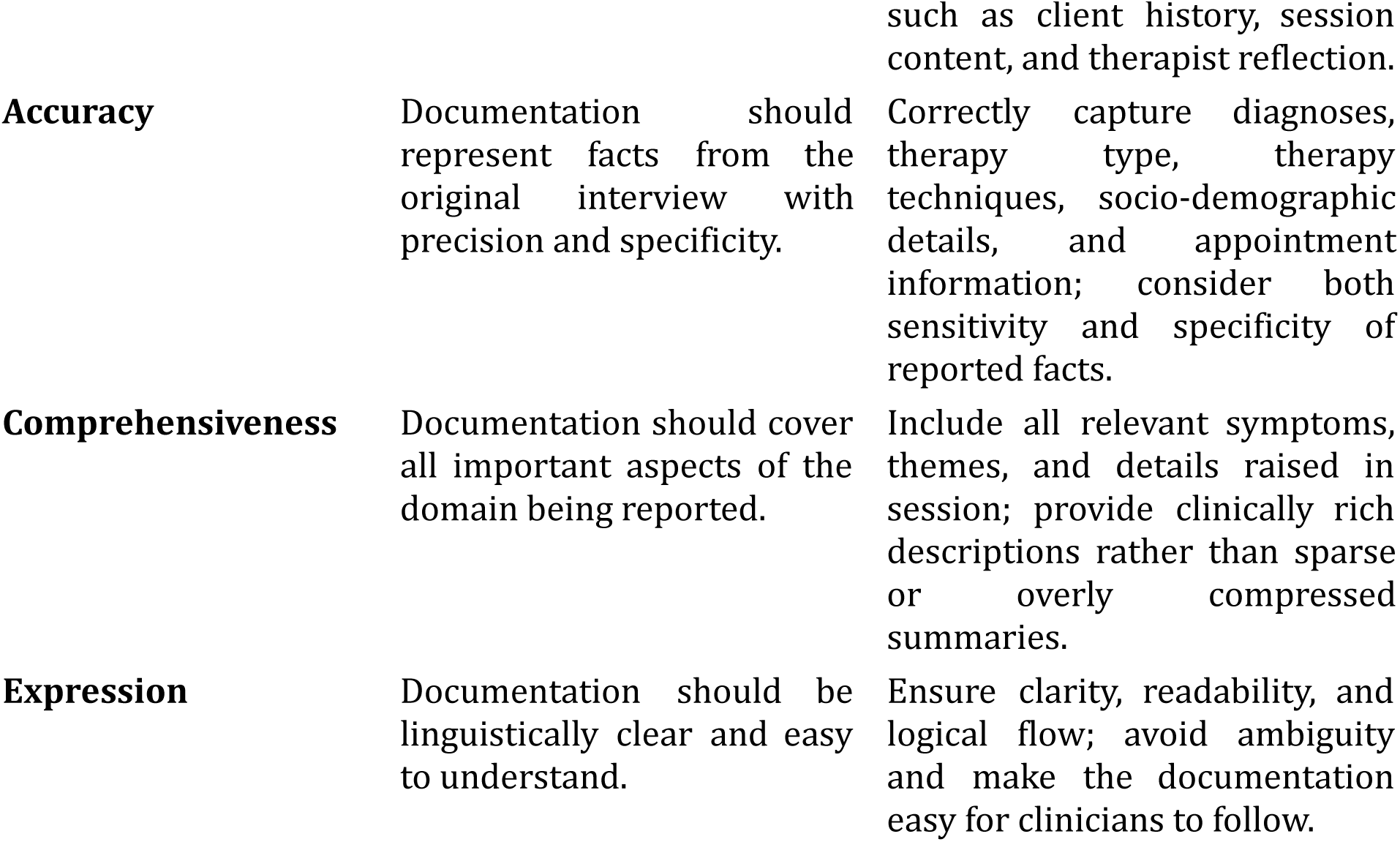
TRACE, a five-domain evaluation framework for AI-assisted documentation in psychotherapy.

During evaluation, each evaluator was provided with sets of iHOPE transcripts along with four corresponding summaries in the iCARE format, through an online interface for rating. Each of these sets of four, included three best LLM generated responses (in synthetic benchmarks) and one human written gold standard summary. The four responses were presented in random order during rating and evaluators were blinded to the model/human identity.

### Statistical Analysis

Scores were summarised across models for each of the five TRACE domains. The unit of analysis was a single rating instance, defined as one evaluator’s rating of one system’s note for one transcript; ratings were pooled without averaging within transcript or within evaluator, and each system contributed 301 rating instances per domain. Summary values were reported as unweighted arithmetic means with standard deviations, with medians, interquartile ranges and mean ranks additionally tabulated in the Supplement. Because every evaluator rated all four summaries derived from the same transcript, the four systems constituted matched rather than independent samples; between-system comparisons were therefore performed using the Friedman test [67] with Nemenyi post-hoc analysis [68,69], with the Kruskal-Wallis test [70] retained as a sensitivity analysis. Because five domain-level tests were performed, omnibus p-values were adjusted using the Holm-Bonferroni procedure [71]; the Nemenyi procedure controls family-wise error within each domain’s pairwise comparisons. As transcripts were rated by between one and five evaluators, all comparisons were repeated after averaging within the transcript as a robustness check. Inter-rater agreement was quantified using ordinal Krippendorff’s α [72], with 95% confidence intervals obtained by bootstrapping over rated units.

### Ethical Considerations

This study did not involve real patient data. All therapy-dialogue data used in this research were simulated conversations derived from publicly available YouTube videos, and did not contain any identifiable, private, or real patient information. As no human subjects’ private or personal data were collected, accessed, or analyzed, formal ethics review board (IRB/REB) approval was not sought, consistent with institutional and regional policies exempting research using publicly available, non-identifiable data from ethics board review. No application or case number was obtained, as no submission was made to an ethics review board.

The mental health professionals who prepared the gold-standard notes and who took part in the blinded expert evaluation were in-house members of the research team, and received no additional compensation for their participation in this study.

This research included local researchers throughout all stages of the research process. Roles and responsibilities were mutually agreed upon by all collaborators prior to the commencement of the study. No health, safety, security, or other risks were posed to participants or researchers during this research.

## Results

### The iCARE Format for Psychotherapy Documentation

An initial review of psychotherapy documentation guidelines identified 25 candidate components, encompassing all domains across different stages of psychotherapy. Following iterative refinement with expert feedback, several items were merged or removed, resulting in the final iCARE framework comprising 17 sections (Table 2). The first iCARE component captures identifying information related to the patient, therapist, treatment setting, and key therapy and session details. In high-resource settings with well-established hospital information systems, such information is generally routinely available. Unfortunately, in resource-limited settings, where digital infrastructure is rudimentary, a substantial proportion of manual documentation time is spent recording these basic but essential details. Additionally, separating all identifying information to a single section helps significantly in ensuring privacy and digital data protection rules compliance. The central CARE components include Chief Concerns and Clinical History, Assessment and Analytical Formulation, Risk identification, Evaluation of progress and action plan. These concepts are familiar to all clinicians and are universal across most clinical history guidelines.

**Table 2.**
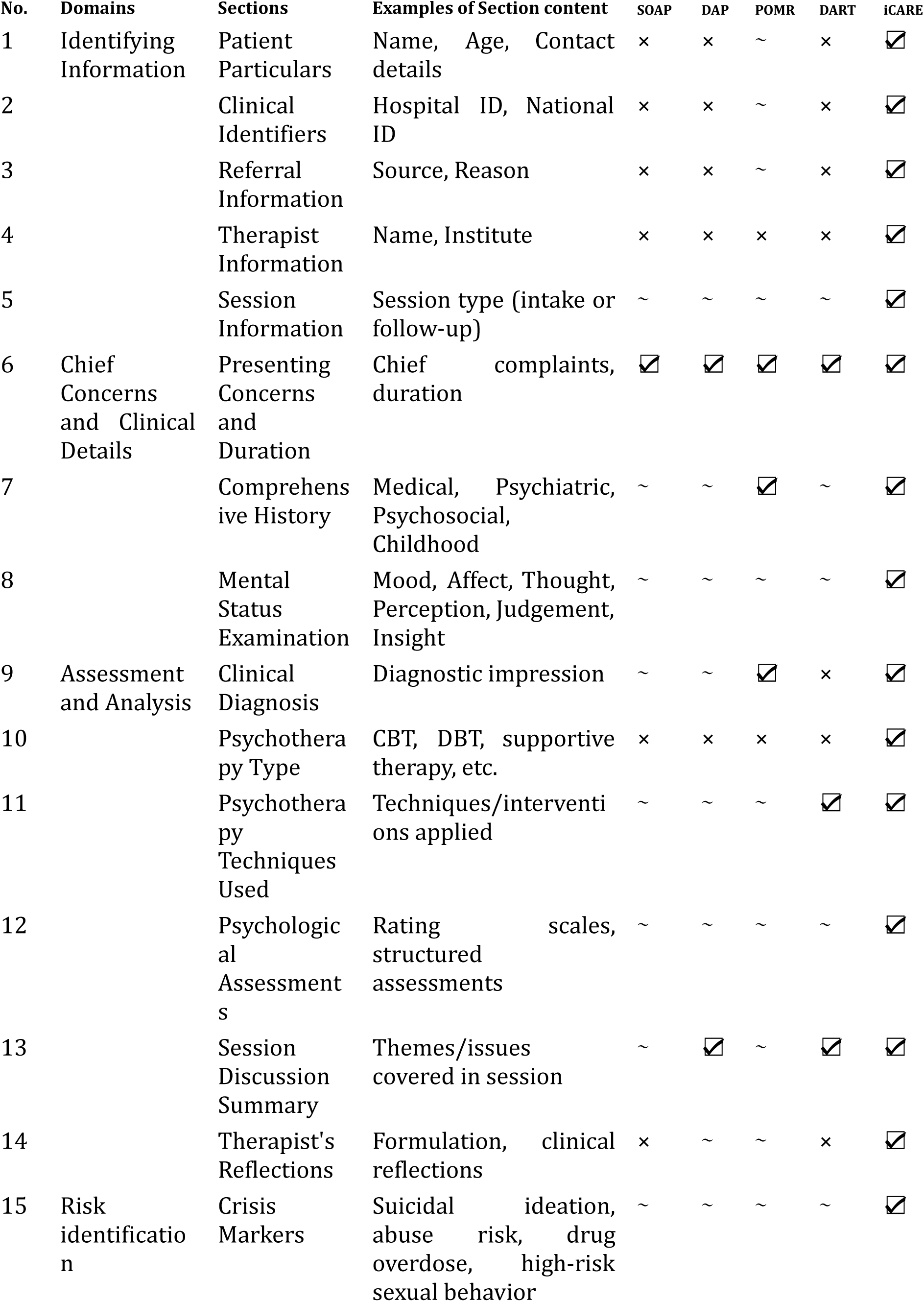
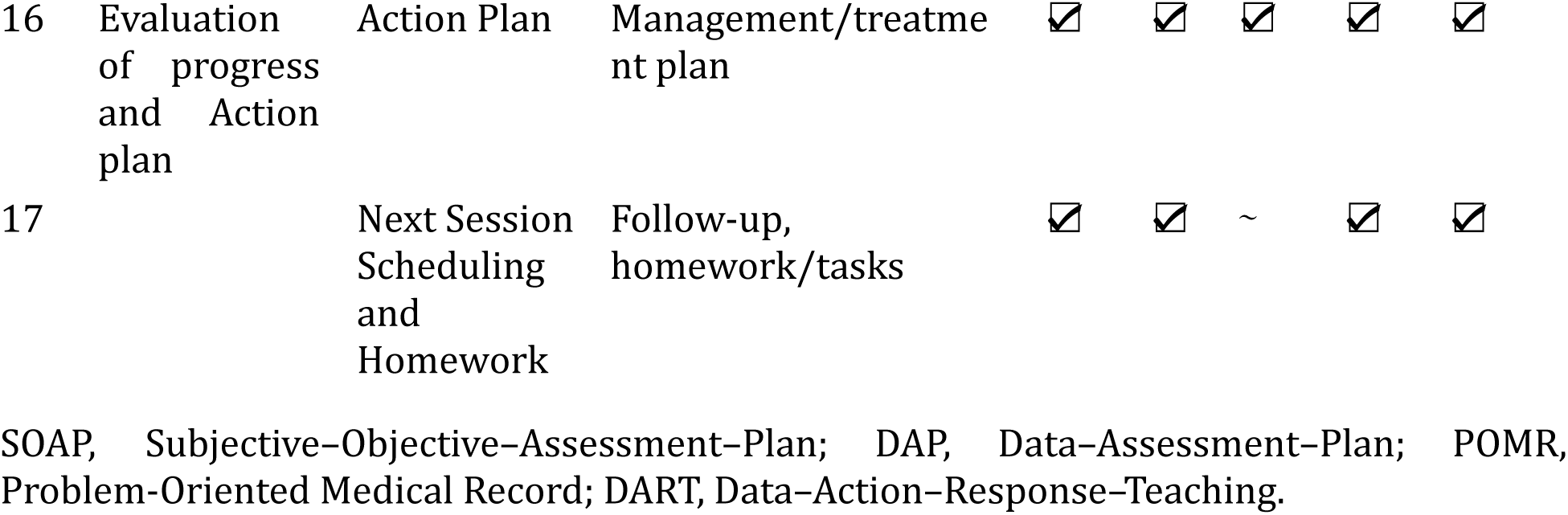
The iCARE documentation format and its comparison with other structured psychotherapy notes.

### The iHOPE Transcript and the iHOPE Dataset

The HOPE+ transcripts, prepared after rigorous manual validation of the original HOPE videos, were used to generate both the human expert summaries (gold standard) as well as the LLM summaries. Multimedia Appendix 1, section S1.4 provides a quantitative overview of iHOPE transcripts. The iHOPE dataset, a corpus of expert generated summaries of the HOPE+ transcript, in the iCARE format, was used as the gold standard against which all LLM models were tested. Multimedia Appendix 1, section S1.5 provides an example of a iHOPE transcript and a corresponding iHOPE summary.

### Benchmarking LLM Performance

We employed automatic evaluation of 11 current LLMs using a combination of lexical metrics (BLEU [53], METEOR [54], ROUGE-L [55]) and semantic metrics (BERTScore [56], BLUERT [57], and InfoLM [58]) to assess model performance comprehensively. Complete details of the 11 LLMs performance on various automatic evaluation metrics, for both zero-shot and one-shot prompting scenarios are available in the Multimedia Appendix 1, sections S1.7 and S1.8. The following sections summarise the major benchmarking findings and their clinical relevance. Figures 2 and 3 provide performance analysis of the top three LLMs (GPT4o-mini, Gemini Pro, and Mistral) across the different benchmarks and aspects of psychotherapy note generation.

I. **Zero-Shot vs One-Shot:** Our overall findings revealed that closed-source models (GPT-4o-mini and Gemini Pro) consistently outperformed open-source models in both zero-shot and one-shot prompting conditions. Among open-source models, Mistral and Orca 2 exhibited relatively better performance. Shifting from zero-shot to one-shot prompting improved model outputs across most of the documentation sections. For instance, GPT-4o-mini’s performance on ‘Clinical Identifiers’ improved from a ROUGE-L of 0.593 to 0.658, and its BERTScore increased from 0.907 to 0.926. A similar trend was observed in most models, although the degree of improvement varied across documentation sections.
II. **Model-specific Observations:** GPT4o-mini demonstrated superior overall performance in most aspects, with the overall highest scores in one-shot settings, and high scores in many zero shot sections. The other top performer, Gemini Pro, in comparison showed dominance in the narrative and interpretative aspects of documentation. Notably, Gemini achieved the highest score in ‘Crisis Markers’ (ROUGE-L: 0.733; BERTScore: 0.912), an important section, requiring high interpretation capability. Furthermore, Gemini Pro often achieved better BLUERT scores than other models, suggesting better fluency and coherence in its generation. For instance, it achieved a positive BLUERT score of 0.298 for ‘Crisis Markers’ while other models achieved negative scores in a one-shot setting. Mistral, an open-source model, gave comparable one-shot performance to GPT4o-mini and Gemini Pro in some sections (e.g.: ‘Patient Particulars’). However, the performance gap widened significantly in more complex aspects of documentation (’Reflections by Therapist’) requiring understanding of the therapeutic process. Among the other open-source models, Orca 2 and Zephyr gave a decent performance in sections which required less synthesis (’Assessments’ and ‘Psychotherapy Type’). Llama 3 and Llama 3.1 demonstrated consistent but moderate performances across the metrics and limited improvement using a one-shot setting. Gemma and MentalLLaMA exhibited the poorest performance with consistently low scores.
III. **Performance across different sections of psychotherapy documentation:** Information-focussed tasks like ‘Patient Particulars,’ ‘Clinical Identifiers,’ and ‘Assessments’ (psychological tests administered) were very well captured by most models, with GPT4o-mini and Gemini Pro consistently achieving top scores. In contrast, temporal-focussed tasks such as ‘Next Session Details’, ‘Past Session Information’ and ‘History’ posed challenges to most models. To illustrate, both GPT4o-mini and Gemini Pro achieved a BLEU score of zero for the sections of ‘Past Session Information’ and ‘Next Session Details’. These results suggest that models handle factual information better than temporal or contextual reasoning. The weighted scores in Figure 2 additionally suggest superiority of closed-source models (GPT4o-mini and Gemini Pro) in interpretative tasks while taking in account the common challenge all model architectures face in temporal reasoning tasks.

**Figure 2.**
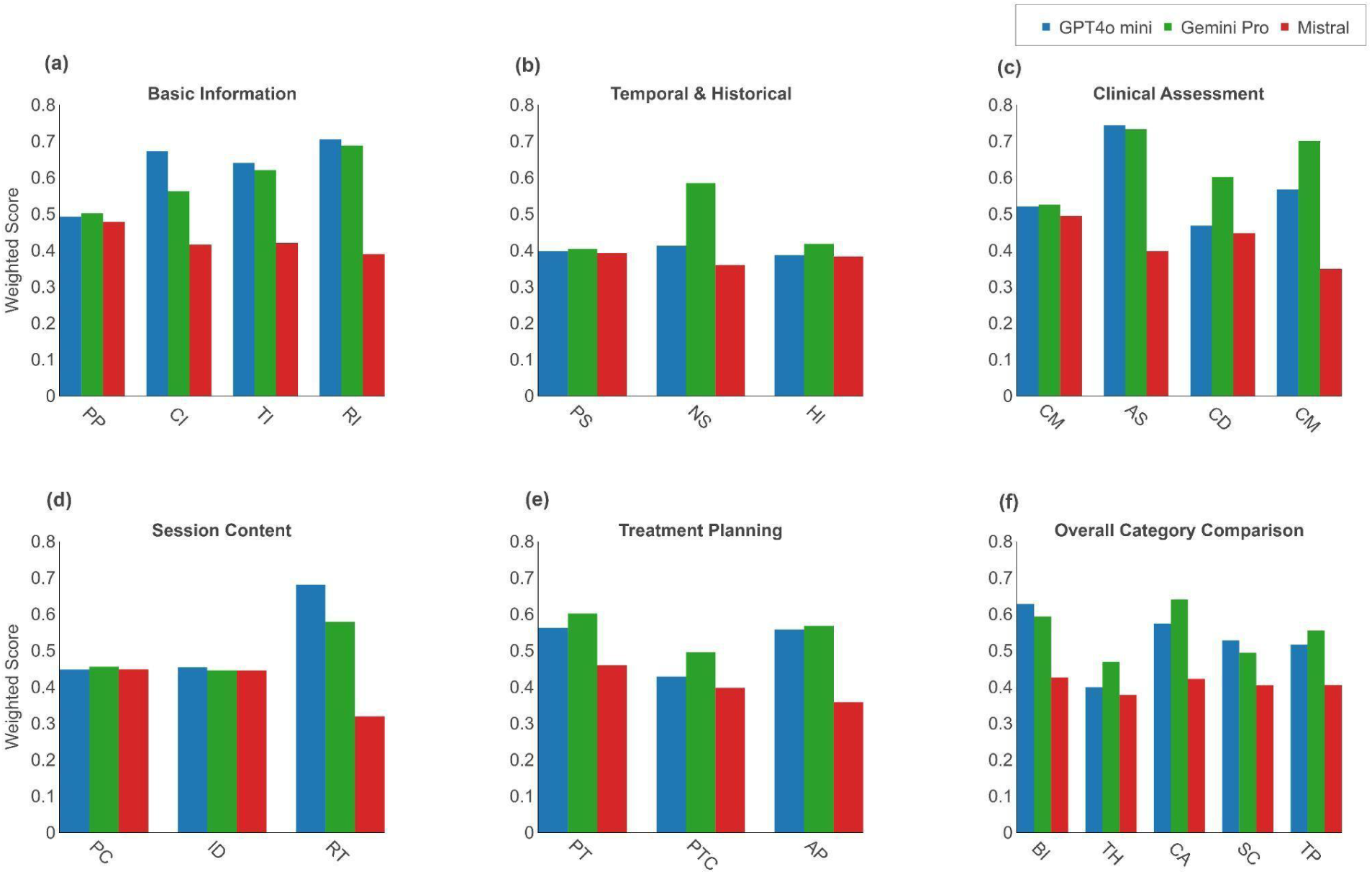
Performance analysis of the top three LLMs (GPT4o-mini, Gemini Pro, and Mistral) across the different aspects of psychotherapy note generation. The first five plots from (a) to (e) show weighted scores (30% METEOR, 30% ROUGE-L, 40% BERTScore) for each aspect within iCARE categories: Basic Information, Temporal & Historical, Clinical Assessment, Session Content, and Treatment Planning. The final plot (f) presents the aggregated performance across all categories to give a holistic overview of the models’ effectiveness in handling different aspects. Note: Each category comprises specific aspects as follows: Information (Patient Particulars (PP), Clinical Identifiers (CI), Therapist Information (TI), Referral Information (RI)); Temporal & Historical (Past Session Information (PS), Next Session Details (NS), History (HI)); Clinical Assessment (Current Mental Status Examination (CMS), Assessments (AS), Clinical Diagnosis by Reviewer (CD),Crisis Markers (CM)); Session Content (Presenting Complaints/Symptoms (PC), Issues Discussed in Current Session (ID), Reflections by Therapist (RT)); and Treatment Planning (Psychotherapy Type (PT), Psychotherapy Technique (PTC), Action Plan (AP)).

**Figure 3.**
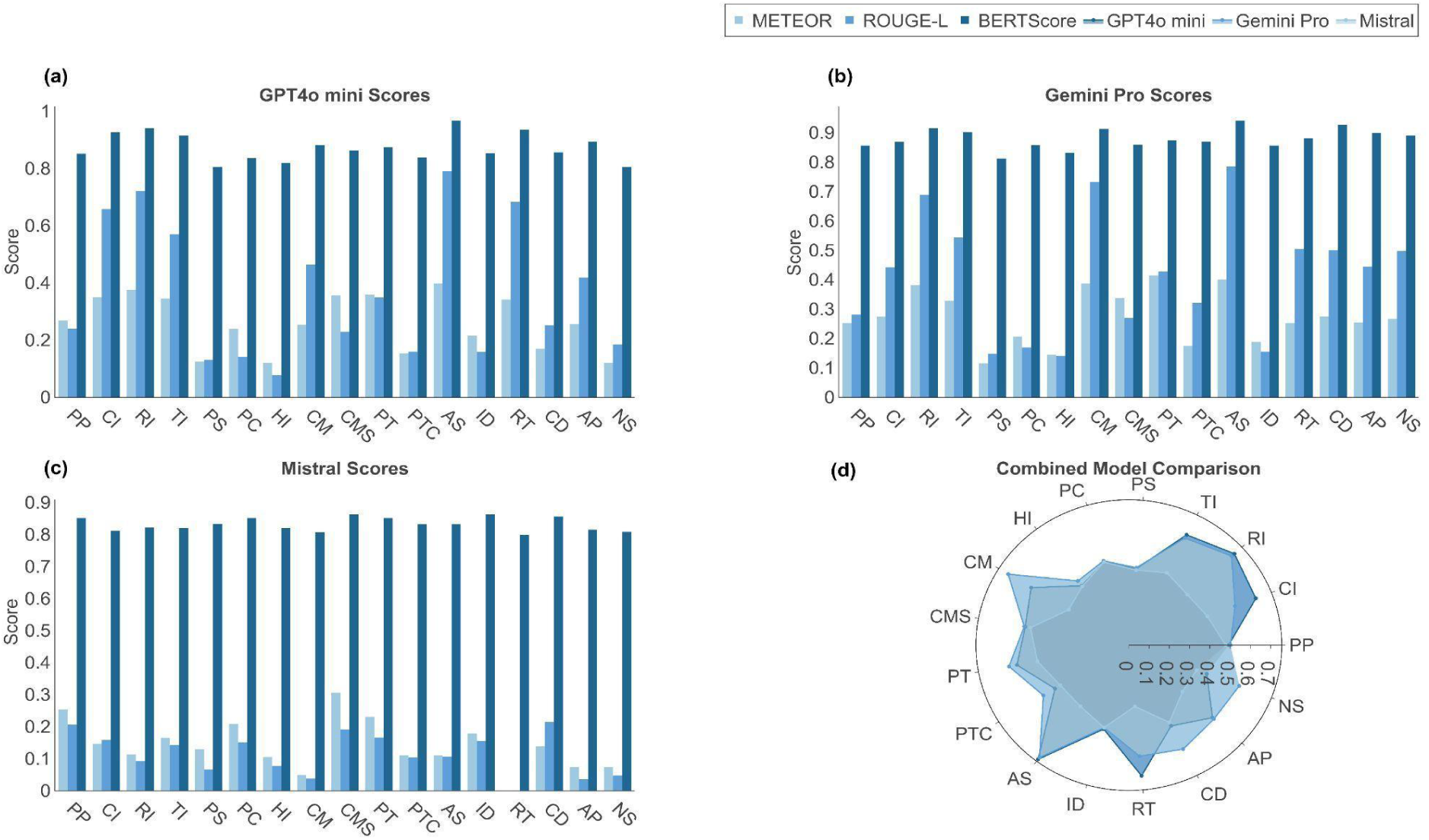
Metric comparison (METEOR, ROUGE-L, and BERTScore) for (a) GPT4o-mini, (b) Gemini Pro, and (c) Mistral across 17 aspects of psychotherapy notes in a one-shot setting. (d) The Radar plot compares the weighted average scores of METEOR, ROUGE-L, and BERTScore of all models across the aspects. Note: Aspects are abbreviated as follows: Patient Particulars (PP), Clinical Identifiers (CI), Referral Information (RI), Therapist Information (TI), Past Session Information (PS), Presenting Complaints/Symptoms (PC), History (HI), Crisis Markers (CM), Current Mental Status Examination (CMS), Psychotherapy Type (PT), Psychotherapy Technique (PTC), Assessments (AS), Issues Discussed in Current Session (ID), Reflections by Therapist (RT), Clinical Diagnosis by Reviewer (CD), Action Plan (AP), and Next Session Details (NS).

### Metric-Specific Observations

We evaluated metrics for lexical similarity and semantic alignment against the reference gold standard documentation developed. All models exhibited low lexical similarity scores, indicating the models’ limitation in achieving exact phrase matches with the reference. This limitation is especially highlighted by a consistently low BLEU score across all aspects in all models. With relatively flexible lexical matching metrics, such as METEOR and ROUGE-L, the models demonstrated a better performance trend. In the case of Gemini Pro, for example, the BLEU score (0.205) was substantially less than the METEOR (0.414) and ROUGE-L (0.427) scores for the documentation aspect ‘Psychotherapy Type’.

Model performance for semantic similarity scores were higher than lexical metric scores across all documentation aspects. BERTScore, in particular, showed strong semantic alignment. For example, in ‘Assessments’, both GPT4o-mini (0.966) and Gemini Pro (0.940) achieved significantly high scores in one-shot. BLUERT scores revealed areas for improvement, particularly in fluency and relevance, with GPT4o-mini scoring -0.511 in ‘Crisis Markers’ documentation. Gemini Pro achieved a relatively better positive BLUERT score of 0.298 for the same aspect. InfoLM scores varied widely for all models, with improvements observed in zero-shot to one-shot settings. These multi-metric analyses suggest that although current models struggle with exact phrase matching of human-written notes, they do retain the text’s underlying semantic meaning and clinical significance of the content.

### Human Evaluation of LLM Performance

Six mental health professionals (3 psychiatrists and 3 psychologists), with clinical experience ranging from 4 to 18 years, (Multimedia Appendix 1, section S1.6) participated in the blinded review, each rating a set of four summaries: three best-performing LLM outputs and one human-written gold-standard note, presented in random order and blinded to source, across the five TRACE domains on a 1-5 Likert scale. Each system, therefore, accrued 301 rating instances per domain across 168 transcripts, and the values in Table 4 are unweighted means of those raw ratings. The results show that Mistral was the highest-scoring LLM on all five TRACE domains, although the human-written notes received the highest scores overall (mean across domains 3.02, versus 2.85 for Mistral, 2.74 for Gemini Pro and 2.72 for GPT-4o-mini). This ordering diverges from the automatic benchmarks (Table 3), in which GPT-4o-mini ranked first overall. Mistral was also the only system to surpass the human-written notes on any domain, scoring marginally higher on Comprehensiveness (2.88 vs 2.83). Because each evaluator rated all four summaries of the same transcript, between-system differences were assessed with the Friedman test for matched samples (n=296 complete quadruples): differences were significant on Trustworthiness (χ²=18.39, df=3, p=7.3×10⁻⁴), Relevance (χ²=55.75, df=3, p=2.4×10⁻¹¹), Accuracy (χ²=34.54, df=3, p=6.1×10⁻⁷) and Expression (χ²=34.43, df=3, p=6.1×10⁻⁷), but not on Comprehensiveness (χ²=3.3, df=3, p=0.348); all p-values are Holm-corrected across the five domains. The same pattern held for the pooled overall score (χ²=19.19, df=3, p=2.5×10⁻⁴). In Nemenyi post-hoc tests, the human notes scored significantly higher than all three LLMs on Relevance (all p<0.001) and Expression (p<0.001, p=0.002 and p=0.002 for GPT-4o-mini, Gemini Pro and Mistral respectively), and significantly higher than GPT-4o-mini and Gemini Pro, but not Mistral, on Trustworthiness (p=0.009, p=0.020 and p=0.383) and Accuracy (p=0.001, p<0.001 and p=0.330). On the pooled overall score Mistral was the only system not significantly distinguishable from the human notes (p=0.079). No pair of systems differed significantly on Comprehensiveness. The only significant difference between two LLMs was Mistral over Gemini Pro on Accuracy (p=0.018); the three models were otherwise statistically indistinguishable from one another. Repeating the analysis after averaging ratings within the transcript left the ordering of systems unchanged. Full test statistics, pairwise p-value matrices, descriptive statistics and a Kruskal–Wallis sensitivity analysis are reported in Multimedia Appendix 1, section S1.9. Inter-rater agreement was low (ordinal Krippendorff’s α=0.10, 95% CI [0.01, 0.18] for the pooled score; 0.04-0.19 across individual domains), reflecting the inherent subjectivity of psychotherapy-note evaluation.

**Table 3.**
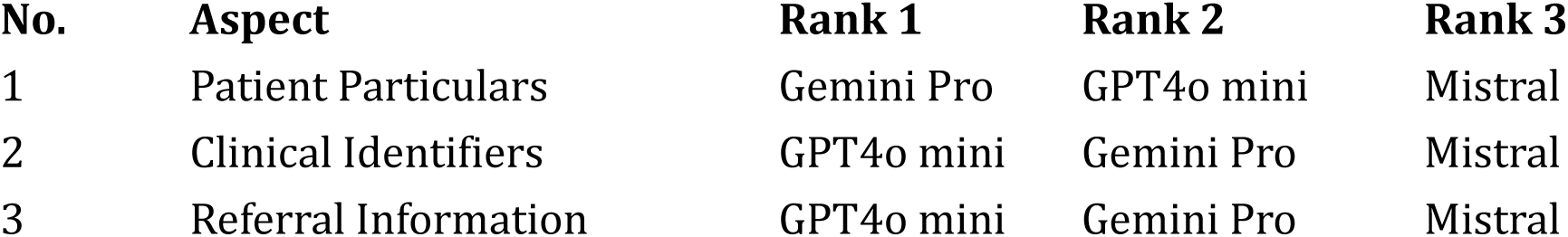
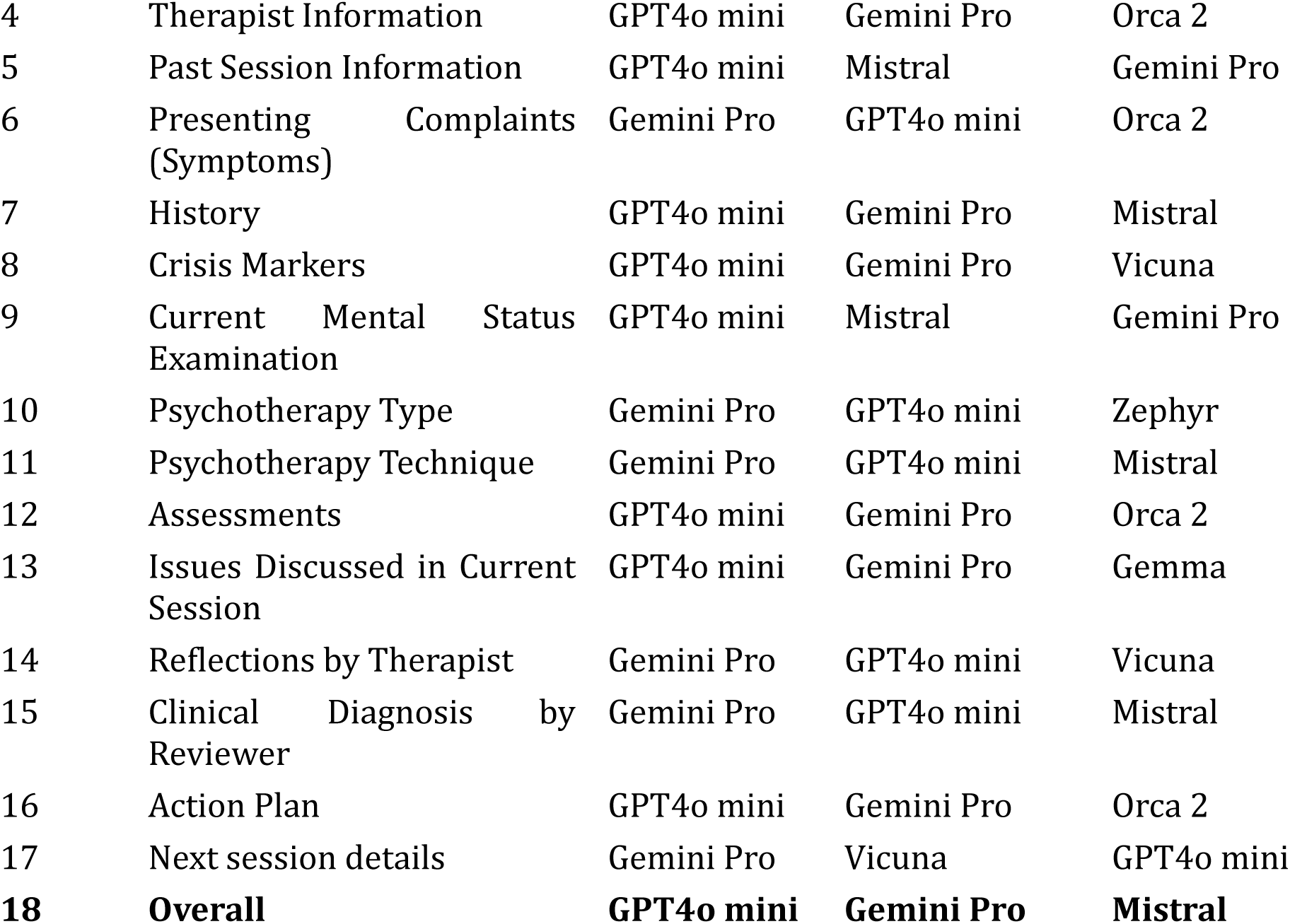
Summary ranking of all models across the 17 domains of iCARE documentation format. Model rankings are based on aggregated scores of BLEU, METEOR, ROUGE-L, BERTScore, BLEURT and InfoLM in one-shot prompting.

| No. | Aspect | Rank 1 | Rank 2 | Rank 3 |
| --- | --- | --- | --- | --- |
| 1 | Patient Particulars | Gemini Pro | GPT4o mini | Mistral |
| 2 | Clinical Identifiers | GPT4o mini | Gemini Pro | Mistral |
| 3 | Referral Information | GPT4o mini | Gemini Pro | Mistral |
| 4 | Therapist Information | GPT4o mini | Gemini Pro | Orca 2 |
| 5 | Past Session Information | GPT4o mini | Mistral | Gemini Pro |
| 6 | Presenting Complaints (Symptoms) | Gemini Pro | GPT4o mini | Orca 2 |
| 7 | History | GPT4o mini | Gemini Pro | Mistral |
| 8 | Crisis Markers | GPT4o mini | Gemini Pro | Vicuna |
| 9 | Current Mental Status Examination | GPT4o mini | Mistral | Gemini Pro |
| 10 | Psychotherapy Type | Gemini Pro | GPT4o mini | Zephyr |
| 11 | Psychotherapy Technique | Gemini Pro | GPT4o mini | Mistral |
| 12 | Assessments | GPT4o mini | Gemini Pro | Orca 2 |
| 13 | Issues Discussed in Current Session | GPT4o mini | Gemini Pro | Gemma |
| 14 | Reflections by Therapist | Gemini Pro | GPT4o mini | Vicuna |
| 15 | Clinical Diagnosis by Reviewer | Gemini Pro | GPT4o mini | Mistral |
| 16 | Action Plan | GPT4o mini | Gemini Pro | Orca 2 |
| 17 | Next session details | Gemini Pro | Vicuna | GPT4o mini |
| **18** | **Overall** | **GPT4o mini** | **Gemini Pro** | **Mistral** |

**Table 4.** Results of expert evaluation of the top three LLM models (GPT4o-mini, Gemini Pro, and Mistral) based on the TRACE evaluation framework. Cells are mean (SD) of raw 1-5 Likert ratings, pooled over 301 rating instances per system per domain, where one rating instance is one evaluator’s rating of one system’s note for one transcript (six evaluators, 168 of the 174 iHOPE sessions; see Methods). Higher is better; ‘Overall’ is the mean of a rating instance’s five domain scores, and the best-performing LLM in each domain is shown in bold. Between-system differences were tested with the Friedman test for matched samples (n=296 complete quadruples) with Nemenyi post-hoc comparisons, Holm-corrected across the five domains; all domains differed significantly except Comprehensiveness. The human notes scored above all three LLMs on Relevance and Expression, and above GPT-4o-mini and Gemini Pro but not Mistral on Trustworthiness and Accuracy; Mistral was not significantly distinguishable from the human notes on the overall score (p=0.079). Medians, IQRs, mean ranks, full test statistics and a Kruskal-Wallis sensitivity analysis are given in Multimedia Appendix 1, section S1.9.

| Model | Trustworthiness | Relevance | Accuracy | Comprehen<br>sive-ness | Expression |
| --- | --- | --- | --- | --- | --- |
| Gemini Pro | 2.78 (0.83) | 2.65 (0.94) | 2.60 (0.85) | 2.80 (0.90) | 2.88 (0.97) |
| GPT-4o mini | 2.76 (0.91) | 2.58 (0.95) | 2.67 (0.95) | 2.78 (0.90) | 2.78 (0.96) |

| Model | Trustworthiness | Relevance | Accuracy | Comprehensiveness | Expression |
| --- | --- | --- | --- | --- | --- |
| **Mistral** | **2.88 (0.88)** | **2.71 (0.86)** | **2.88 (0.87)** | **2.88 (0.85)** | **2.90 (0.87)** |
| Human | 3.01 (0.97) | 3.09 (0.88) | 2.99 (0.93) | 2.83 (0.95) | 3.19 (0.90) |

## Discussion

### Principal Findings

This study makes four core contributions to the field of AI in psychotherapy.

First, we developed iCARE, a structured and clinically grounded framework for AI documentation of psychotherapy notes. Unlike conventional note formats such as SOAP and DAP, which were developed to reduce the administrative burden on human clinicians [73], iCARE was designed from first principles around the core clinical needs of the therapist in a post-AI world. Our review of AI-scribing solutions for therapy sessions [16, 24, 25, 74] indicate that most models provide summaries which are too brief for meaningful clinical use beyond administrative documentation. Our work emerges from a deep collaboration among AI researchers, psychiatrists, and psychologists, and rethinks psychotherapy documentation for a Therapist-AI collaborative future.

Second, we re-transcribed, speaker-diarized, and manually refined the HOPE dataset, to form the accurate iHOPE transcript, and generated expert-written therapy notes in the iCARE format (iHOPE dataset). This human “gold standard” reference set can be used to train and test AI systems attempting to automate mental health documentation.

Third, our extensive benchmarking of various LLMs provides valuable insights into the current state of AI capabilities in mental health documentation. Evaluation across multiple metrics and aspects of documentation offer a nuanced understanding of the models’ strengths and limitations. Models exhibited domain-specific strengths, with different models excelling at different aspects of clinical notes, thereby raising the interesting possibility of developing multi-agent systems for better documentation. The tested LLMs also faced persistent challenges in handling the past session information and future action plans, suggesting fundamental limitations in their temporal reasoning. Furthermore, their low performance in identification of facts of high clinical relevance (eg: drug use), limit their clinical trustworthiness. We additionally conducted extensive human clinical evaluation of our model outputs by a team of mental health experts. A 2026 systematic review [64] on the topic highlights the need for such clinical evaluation and mentions the lack of such, a shortcoming of most current research. Our expert evaluation strengthens our results and generates the interesting finding that clinical preference might not always match with synthetic benchmarking performance. In our blinded review the open-source Mistral model, which ranked third overall on the automatic benchmarks, was the highest-rated LLM on all five TRACE domains and the only system to exceed the human notes on any domain.

Fourth and finally, we developed a new benchmarking framework (TRACE) for evaluating mental health related documentation, prioritizing models’ trustworthiness and safety, as well as its capacity for analytical and reflective reasoning. Our evaluation metric aligns with the specific needs of the subject, which go beyond the already rigorous requirements of any medical documentation.

### Limitations

In the rapidly changing field of LLMs, all benchmarks lose relevance with time as newer models and versions supersede capabilities of their forebearers. Our need to choose LLM models early in the process arose from the stepwise nature of this study and is a limitation faced by most researchers in this field. However, our aim in this work was not merely model benchmarking. Rather, we tried to understand the needs of the clinicians, through deep interdisciplinary alignment, and attempted a new paradigm for AI-psychotherapy documentation. The iCARE note format, iHOPE summaries and the TRACE evaluation framework developed along the way, will help all future research exploring the interface of AI and mental healthcare. Second, as our work is based on publicly available therapy sessions in the English language, it may not capture diversity found in multilingual settings. Finally, our data-source consisted mostly of short to medium length transcripts. Hence, performance on hour-long (average therapy session length) therapy transcripts or over longitudinal multiple therapy sessions, needs to be explored, particularly in the light of our own finding, where most models fared poorly in timeline understanding. As a major part of human communication is non-verbal, future research should also explore multimodal datasets like video sessions and include voice or other nonverbal cues to enhance the depth of the automated documentation.

### Conclusions

To conclude, our work contributes meaningfully to the evolving field of AI and mental health, particularly by developing a framework for automated psychotherapy documentation, while maintaining clinical validity and therapeutic value. We emphasise that while AI can significantly aid in clinician facing tasks, the complex nature of psychotherapy requires the experts’ careful consideration, before AI is used in any client facing task. We, in this paper, therefore, endeavour to harness the power of LLMs to assist clinicians and ease the clerical burden of therapists and not to replace them through automated systems. Future research should explore similar task-sharing and task-shifting opportunities in mental healthcare, that are safe for the client and beneficial to the clinician, to bridge the huge gap between population need and available services.

## Acknowledgments

The authors would like to express their sincere gratitude to the following Psychiatrists and Clinical Psychologists for their valuable time, expertise, and thoughtful evaluations that greatly contributed to the quality and rigor of this work: Aishwarya Raj, Akanksha, Anurag Sharma, Anusha Thakur, Ayushi Sobhani, Farhin Mustaque, Jemima Jacob, M. Kaur, Manish Shukla, Medhavi Sood, Neha Singh, Nitish Kumar, Ojasvi Meena, Paakhi Srivastava, Pankhuri Soni, R. K. Meena, Ratnakar, Rishabh Jain, Shireen Chaturvedi, Shreyanshi Sata, Sonia Puar, Sumit Bakshi, Vidushi Sharma.

Generative artificial intelligence was not used to draft, write or generate any part of this manuscript. Grammarly, a language-editing tool, was used on author-written text to check grammar and improve readability. The large language models evaluated in this work are the object of study and are described in the Methods; they were not used in the preparation of the manuscript.

## Funding

Tanmoy Chakraborty acknowledges the support of the Rajiv Khemani Young Faculty Chair Professorship in Artificial Intelligence. Koushik Sinha Deb and Tanmoy Chakraborty acknowledge the support of Indian Council of Medical Research for the projects IIRPIG-2025-01-01778 and IIRPSG-2025-01-03706.

## Conflicts of Interest

None declared.

## Data Availability

All conversation transcripts used in this study and all source code for benchmarking experiments are publicly available in https://github.com/ai4mhx/iCARE (accessed April 26, 2025) [75].

## Authors’ Contributions

*PKA and TC conceptualized the study. PKA and SahS developed the methodology and software, and carried out the formal analysis and investigation. PKA, SMS were responsible for data curation and visualization. SurS, RC, CS, PC, SKG, and KSD developed the iCARE documentation format through iterative clinical review, prepared the gold-standard session notes, and conducted the blinded expert evaluation. KSD and SKG designed and supervised the human evaluation framework. TC and KSD acquired funding, provided resources, and supervised the project. PKA and KSD wrote the original draft. All authors validated the results, contributed to writing, review and editing, and approved the final manuscript*.

## Abbreviations

AI: artificial intelligence
BERTScore: bidirectional encoder representations from transformers score
BLEU: bilingual evaluation understudy
BLEURT: bilingual evaluation understudy with representations from transformers
DAP: data, assessment, plan
iCARE: identifying information, chief concerns and clinical history, assessment and analysis, risk identification, evaluation of progress and action plan
iHOPE: improved health-oriented psychotherapeutic evaluation corpus
InfoLM: information-theoretic metric for language model evaluation
LLM: large language model
METEOR: metric for evaluation of translation with explicit ordering
ROUGE-L: recall-oriented understudy for gisting evaluation, longest common subsequence
SOAP: subjective, objective, assessment, plan
TRACE: trustworthiness, relevance, accuracy, comprehensiveness, expression

## Multimedia Appendices

**Multimedia Appendix 1**. Supplementary information: instruction prompts for each iCARE section, descriptions of the large language models tested, descriptions of the automatic evaluation metrics, session- and utterance-level statistics of the iHOPE transcripts, an example iHOPE transcript and structured summary, details of the expert evaluators, full zero-shot and one-shot benchmarking results, and the full statistical comparison of expert TRACE ratings.

**Multimedia Appendix 2**. AI-assisted documentation in psychotherapy: characteristics, technology and findings of prior studies.

## Notes

### Competing Interest Statement

The authors have declared no competing interest.

### Summary of Updates

This version of the manuscript has been revised to update the language throughout the manuscript, strengthen the statistical methodology, and improve the evaluation methodology and reporting.

