## Supplementary Document 1 for "Clinically Grounded AI-Scribing in Psychotherapy: Benchmarking LLMs Against Expert Documentation in the iCARE Framework"

Multimedia Appendix 1. Supplementary information

This appendix contains the instruction prompts used for each iCARE section, descriptions of the large language models tested and of the automatic evaluation metrics, session- and utterance-level statistics of the iHOPE transcripts, an example iHOPE transcript with its structured summary, details of the expert evaluators, the full zero-shot and one-shot benchmarking results, and the full statistical comparison of the expert TRACE ratings.

Abbreviations used in this appendix: AI (artificial intelligence); BERTScore (bidirectional encoder representations from transformers score); BLEU (bilingual evaluation understudy); BLEURT (bilingual evaluation understudy with representations from transformers); iCARE (identifying information, chief concerns and clinical history, assessment and analysis, risk identification, evaluation of progress and action plan); iHOPE (improved health-oriented psychotherapeutic evaluation corpus); InfoLM (information-theoretic metric for language model evaluation); IQR (interquartile range); LLM (large language model); METEOR (metric for evaluation of translation with explicit ordering); ROUGE-L (recall-oriented understudy for gisting evaluation, longest common subsequence); SD (standard deviation); TRACE (trustworthiness, relevance, accuracy, comprehensiveness, expression).

### **S1.1. Instruction Prompts**

Table S1 presents the instructional prompts for summarizing notes based on definitions of each aspect. These prompts were created in collaboration with mental health experts.

| **Aspect(s)** | **Instruction(s)** |
| --- | --- |
| **Patient Particulars** | Extract the following patient particulars from the given mental health dialogue: Name, Age, Sex, Marital Status, Education, Residence, Phone, Mobile, Email, and details of any caregivers or family members present. Record each piece of information in the format Field: Value and separate each dataset with a semicolon. For example: Name: Mr. X; Mobile: +91 XXXXXXXXX. |
| **Clinical Identifiers** | From the given mental health dialogue, identify and extract clinical identifiers related to the clinical setting where the patient is being seen. Include details such as Hospital Name, Hospital ID, OPD/Inpatient/Telepsychiatry/Clinic, Room Number, Bed Number, Date of Assessment, and other relevant clinical information. Record each piece of information in Field: Value format, separated by a colon, and each dataset with a semicolon. For example, Setting: OPD; UHID: AIIMS-12234; Date: dd/mm/yyyy. |
| **Referral Information** | From the provided mental health dialogue, extract any referral information related to the clinician or clinical establishment where the patient was previously seen. Include details such as the Doctor’s Name, Previous Hospital Name, Clinical Department, School, Office, Legal, Administrative, or other relevant settings. Additionally, specify the reason for referral in a separate entry. Record each piece of information in Field: Value format, separated by a colon, and each dataset with a semicolon. For example: Referred by: Psychiatrist; Referrer Name: Dr. Amit; Referring speciality: Cardiology; Reason for Referral: Panic attacks |
| **Therapist Information** | From the given mental health dialogue, identify and extract information about the clinician or clinical establishment where the patient is being seen. Include details such as Doctor’s Name, Current Hospital Name, Clinical Department, and other relevant information. Record each piece of information in Field: Value format, separated by a colon, and each dataset with a semicolon. For example, the hospital’s name is Dr. Smith; the current hospital is City Hospital; and the clinical department is Psychiatry. |
| **Past Session Information** | From the given mental health dialogue, identify and extract information about the clinician or clinical establishment where the patient is being seen. Include details such as Doctor’s Name, Current Hospital Name, Clinical Department, and other relevant information. Record each piece of information in Field: Value format, separated by a colon, and each dataset with a semicolon. For example, the hospital’s name is Dr. Smith; the current hospital is City Hospital; and the clinical department is Psychiatry. |
| **Presenting Complaints (Symptoms)** | From the given mental health dialogue, extract and document the presenting complaints (symptoms) reported by the patient. Present these symptoms as a numbered list, specifying each symptom and its duration. Distinguish between ‘Presenting Symptoms’ (current issues prompting the visit) and ‘Chief Complaints’ (major symptoms throughout the illness, even if not currently present). If the duration is not available, record it as NA. Separate each symptom entry with a semicolon. For example: 1. Panic attacks: 3 years; 2. Low mood: 2 months. |
| **History** | From the provided mental health dialogue, extract and summarize the patient’s history into a coherent paragraph, focusing on the following sections: Present History, Negative History, Past History, Medical History, Substance Use History, Family History, and Personal History. Document each section in the format: Section: Summary, separating each section with a semicolon. Include only the sections that are present in the dialogue. For example, Present History: Acute anxiety for 3 months; Medical History: Hypertension. |
| **Crisis Markers** | From the given mental health dialogue, extract and document any crisis markers related to the risk of harm to self or others. Include details such as Harm to Self: Wish to die, suicidal ideations (including duration, frequency, severity, intrusive, coping strategies), suicide attempts (including lethality and suicidality), refusal or inability to eat and drink, and non-suicidal self-injurious behaviour (NSSI). Harm to Others: Agitation, violence, homicidal. Other Risk Factors: Family history of suicide, high-risk sexual behaviour, high-risk substance use (e.g., IV drug use), and legal crises. |
| **Current Mental Status Examination** | From the provided mental health dialogue, extract and summarize the patient’s current Mental Status Examination (MSE) in the following structured format: General Appearance & Behavior, Speech, Mood & Affect, Thought, Perception, Cognitive Functions, Judgment and Insight. Document only the positive findings, omitting any absent findings. Each section should be in the format: Section: Details, separated by a semicolon. For example, Thought: Preoccupation, hopelessness; Speech: Increased rate; Mood: Depressed. |
| **Psychotherapy Type** | From the provided mental health dialogue, determine and document the type and mode of psychotherapy used. Identify the school or type of therapy mentioned or implied in the dialogue, such as Dynamic (Supportive/Analytical), Cognitive, Behavioral, ERP, etc. Also, specify the mode of therapy, such as Individual, Couple, Group, or Family. Use the format: Type: Therapy Type; Mode: Therapy Mode, separating each entry with a semicolon. For example: Type: CBT; Mode: Individual |
| **Psychotherapy Technique** | From the provided mental health dialogue, extract and document the specific psychotherapy techniques used in the session. Look for mentions of techniques such as Psychoeducation, Reflective Listening, Behavioral Activation, Journaling, Supportive Techniques, Thought Challenging, Cognitive Restructuring, Problem Solving, Analytical Techniques, or any other relevant methods. If multiple techniques are mentioned, list them in a comma-separated format. Examples of these are psychoeducation, reflective listening, and cognitive restructuring. |
| **Assessments** | From the provided mental health dialogue, extract and document any assessments or scales used during the session. If the specific name of the instrument is men- tioned, include it; otherwise, summarize the purpose of the assessment, such as IQ, Personality, Disability, Projective, etc. Use the format Instrument Name or Pur- pose, separating each entry with a semicolon. For example, the Beck Depression Inventory and Personality Assessment. |
| **Issues Discussed in the Current Session** | From the provided mental health dialogue, extract and list the issues discussed during the current session. Document each issue sequentially. For example: 1. Anxiety about work performance; 2. Difficulty in maintaining personal relationships; 3. Lack of motivation. |
| **Reflections by Therapist** | From the provided mental health dialogue, extract any reflections made by the therapist after the session. These reflections should capture the therapist’s subjective understanding of the therapy interaction, including observations on the client’s demeanour, attitude towards the therapist, rapport,trust, transference, receptiveness to therapy techniques, and attitude towards homework. Note that while not all therapy types require these reflections, they are a common component in dynamic therapy schools. |
| **Clinical Diagnosis by Reviewer** | From the provided mental health dialogue, extract and document the clinical diagnoses made by the reviewer. List each diagnosis separated by a semicolon. For example, Anxiety, Fear, and OCD. |
| **Action Plan** | From the provided mental health dialogue, extract and document the action plan recommended for the client. This includes any techniques, behaviours, or homework assignments the client has been asked to undertake following the session. |
| **Next session details** | From the provided mental health dialogue, extract and document the details for the next session. This includes the date, place, and time of the session. Additionally, note if the patient is required to bring anyone with them. |

### **S1.2. Description of the LLM models tested**

| **Model** | **Number of Parameters** | **Context length** |
| --- | --- | --- |
| Goggle/gemma-7b-it[1] | 7B | 8K |
| Huggingfaceh4/zephyr-7b-beta[8] | 7B | 32K |
| Klyang/mentallama-chat-7B[3] | 7B | 4K |
| Lmsys/vicuna-7b-v1.5[7] | 7B | 4K |
| Meta-llama/Llama-3.1-8B-Instruct[2] | 8B | 128k |
| Meta-llama/Llama-3-8B-Instruct[2] | 8B | 8k |
| Microsoft/Orca-2-7b[5] | 7B | 4K |
| Microsoft/Phi-3-small-8k-instruct[6] | 7B | 8k |
| Mistralai/Mistral-7B-Instruct-v0.2[4] | 7B | 32k |
| Gemini pro[10]^#^ | - | 128K |
| GPT4o-mini[9]^#^ | - | 128K |

^#^denotes the closed-source models

### **S1.3. Automatic Evaluation Metrics**

| **Metric** | **Full form** | **Main focus** | **How it works** | **Strengths** | **Limitations** |
| --- | --- | --- | --- | --- | --- |
| **BLEU** | **Bilingual Evaluation Understudy** | Precision of n-gram overlap | Measures how many n-grams in the generated summary also appear in the reference summary; includes a brevity penalty | Simple, widely used, easy to compute | Does not capture recall well; weak for semantic equivalence |
| **ROUGE** | **Recall-Oriented Understudy for Gisting Evaluation** | Recall of overlap | Compares overlap between generated & reference summaries; **ROUGE-N** uses n-grams, **ROUGE-L** uses longest common subsequence | Standard for summarization; good for content coverage | Relies mainly on lexical overlap; limited semantic understanding |
| **METEOR** | **Metric for Evaluation of Translation with Explicit Ordering** | Precision + recall, with more emphasis on recall | Aligns generated and reference text using exact matches, stemming, synonyms, and word order; uses WordNet | Better correlation with human judgment than BLEU in many tasks; handles synonymy | Still partly surface-form based; depends on external lexical resources |
| **BERTScore** | **Bidirectional Encoder Representations from Transformers Score** | Semantic similarity | Uses BERT contextual embeddings to compare words in generated and reference summaries through cosine similarity, then aggregates precision, recall, and F1 | Captures semantic similarity beyond exact word overlap | More computationally expensive; depends on embedding model quality |
| **BLEURT** | **Bilingual Evaluation Understudy with Representations from Transformers** | Learned similarity aligned with human judgment | Uses a BERT-based model fine-tuned on synthetic data and human ratings to score generated text | Strong correlation with human judgments; robust across text generation tasks | Requires pretrained/fine-tuned model; less transparent than overlap-based metrics |
| **InfoLM** | **-** | Adequacy and fluency using information theory | Uses language model embeddings and information-theoretic measures such as KL divergence to compare generated and reference texts | Captures deeper semantic relationships and content preservation | More complex and less intuitive than traditional metrics |

### **S1.4. Overview of session-level and utterance-level details of the HOPE+ Transcripts**

| **Parameters** | **Value** |
| --- | --- |
| Number of conversations | 174 |
| Total number of utterances | 27,200 |
| Average utterances per conversation | 156.32 |
| Total number of patient utterances | 13,157 |
| Total number of therapist utterances | 14,043 |
| Average patient utterances per conversation | 75.61 |
| Average therapist utterances per conversation | 80.71 |
| Total patient talk-time (seconds) | 36,618.46 |
| Total therapist talk-time (seconds) | 38,624.68 |
| Average patient talk-time per conversation (seconds) | 210.45 |
| Average therapist talk-time per conversation (seconds) | 221.98 |

### **S1.5. Example of iHOPE Transcripts and iHOPE Summary.**

#### **Table S1.5.1. Example iHOPE Transcript Excerpt (abbreviated)**

| **Speaker** | **Transcript Excerpt (HOPE+)** |
| --- | --- |
| **Therapist** | So Bethany, can you tell me what this week has looked like for you? |
| **Bethany** | I'm dead to the world… I'm just closed off in my own little sad universe, I guess. |
| **Therapist** | So you're feeling isolated from your family? |
| **Bethany** | Yeah… it's just been a really tough week with my husband. We've been fighting a lot, as usual. |
| **Therapist** | So you feel that even though you made some progress by looking for jobs this week, you didn't feel much accomplishment from that because your husband didn't acknowledge it. |
| **Bethany** | Yeah, he doesn't appreciate anything I do… it just makes my whole body ache. It just makes me feel empty. |
| **Therapist** | Do you trust me to put it in a little jar, and be able to put all that hurt in a little jar? … Let's try closing your eyes. We'll do some breathing exercises here. |
| **Bethany** | I just feel a lot of guilt… I feel guilty for even feeling hurt, but I feel guilty for feeling good. |
| **Therapist** | So I want you to picture yourself over here. This is the side of you that says you don't deserve to be able to feel guilty and that you don't deserve to be hurt. (Empty-chair technique / thought challenging) |
| **Bethany** | I can feel hurt without feeling guilty… it doesn't make me selfish. I almost feel things feel lighter, like some of that guilt is gone. |
| **Therapist** | So we're actually out of time for today. I want you to take this awareness with you this week, and then we'll pick up again next week. |

#### **Table S1.5.2. iHOPE Structured Summary (abbreviated)**

| **iHOPE Field** | **Summary Detail** |
| --- | --- |
| **Name** | Bethany |
| **Sex** | Female |
| **Age** | Nil |
| **Occupation** | Nil |
| **Referral Source** | Nil |
| **Session Type** | Follow-up session |
| **Presenting Symptoms** | Depressed and anxious |
| **Chief Complaints** | 1) Feels guilty  2) Tries to please family members  3) Hard to acknowledge her own needs  4) Feels sad due to the behavior of her family |
| **Past Psychiatric / Medical History** | Nil |
| **Family History** | Nil |
| **Mental Status Examination (MSE)** | General appearance & behavior: NAD  Speech, Mood & Affect: Depressed  Thought: Negative  Cognitive functions: Slightly impaired  Judgement and Insight: Intact |
| **Therapy Type / Mode** | CBT; Mode: Individual/Behavioral. Techniques: Reflective listening, Thought challenging |
| **Risk Assessment** | Nil |
| **Session Summary (Themes Discussed)** | Problems: Patient feels invalidated when her efforts go unnoticed by her family. She feels frustrated and sad, and guilty for both experiencing hurt and for feeling good.  Techniques: Therapist asked her to visualize and do breathing exercises to temporarily distance herself from her hurt and create mental space to reflect on her feelings. |
| **Substance Use** | Nil |
| **Diagnosis** | Anxiety |
| **Plan / Homework** | The therapist wanted her to take this awareness with her for the week, until the next session. |
| **Follow-up** | Next week |
| **Session Number** | 4 |

### **S1.6. Details of expert evaluators**

| **Name** | **Educational Qualifications** | **Years of Experience** |
| --- | --- | --- |
| Evaluator-1 | MD, Psychiatry | 18 years |
| Evaluator-2 | MD, Psychiatry | 10 years |
| Evaluator-3 | MD, Psychiatry | 5 years |
| Evaluator-4 | MA, M.Phil in Psychology | 14 years |
| Evaluator-5 | MA, M.Phil in Psychology | 4 years |
| Evaluator-6 | MA, M.Phil Psychology | 4 years |

### **S1.7. Zero-shot results across different aspects of psychotherapy note summarization.**

The table reports comprehensive zero-shot results of 11 LLMs using six automatic metrics: BLEU (exact matching), METEOR and ROUGE-L(flexible lexical matching), BERTScore (semantic similarity), BLUERT (fluency and relevance), and InfoLM (semantic distance). The scores demonstrate varying model capabilities across 17 different aspects of psychotherapy notes, with closed-source models (GPT4o mini, Gemini Pro) generally outperforming open-source alternatives.

| **Model(s)** | **Aspect(s)** | **BLEU** | **METEOR** | **ROUGE-L** | **BERT** | **BLEURT** | **InfoLM** |
| --- | --- | --- | --- | --- | --- | --- | --- |
| **GPT4o mini** | Patient Particulars | 0.050 | 0.260 | 0.222 | 0.846 | -1.239 | 2.014 |
|  | Clinical Identifiers | 0.000 | 0.325 | 0.593 | 0.907 | -0.190 | 0.959 |
|  | Referral Information | 0.000 | 0.453 | 0.887 | 0.976 | 0.672 | 0.282 |
|  | Therapist Information | 0.050 | 0.367 | 0.623 | 0.928 | -0.014 | 1.564 |
|  | Past Session Information | 0.000 | 0.134 | 0.123 | 0.806 | -1.737 | 3.099 |
|  | Presenting Complaints (Symptoms) | 0.012 | 0.263 | 0.148 | 0.835 | -0.996 | 2.353 |
|  | History | 0.012 | 0.132 | 0.079 | 0.820 | -1.485 | 2.575 |
|  | Crisis Markers | 0.000 | 0.245 | 0.456 | 0.877 | -0.465 | 1.775 |
|  | Current Mental Status Examination | 0.041 | 0.354 | 0.218 | 0.853 | -0.721 | 2.018 |
|  | Psychotherapy Type | 0.238 | 0.343 | 0.333 | 0.875 | -0.626 | 2.224 |
|  | Psychotherapy Technique | 0.000 | 0.153 | 0.164 | 0.835 | -0.967 | 2.449 |
|  | Assessments | 0.000 | 0.428 | 0.854 | 0.972 | 0.664 | 0.499 |
|  | Issues Discussed in Current Session | 0.024 | 0.218 | 0.171 | 0.851 | -0.556 | 2.352 |
|  | Reflections by Therapist | 0.000 | 0.305 | 0.610 | 0.919 | 0.058 | 1.073 |
|  | Clinical Diagnosis by Reviewer | 0.000 | 0.201 | 0.311 | 0.865 | -0.478 | 2.317 |
|  | Action Plan | 0.004 | 0.218 | 0.363 | 0.883 | -0.571 | 1.385 |
|  | Next Session Details | 0.000 | 0.101 | 0.109 | 0.782 | -1.958 | 2.881 |
| **Gemini Pro** | Patient Particulars | 0.051 | 0.260 | 0.228 | 0.843 | -1.239 | 2.010 |
|  | Clinical Identifiers | 0.000 | 0.204 | 0.256 | 0.822 | -1.334 | 1.795 |
|  | Referral Information | 0.000 | 0.241 | 0.283 | 0.820 | -1.202 | 2.192 |
|  | Therapist Information | 0.025 | 0.251 | 0.304 | 0.854 | -0.917 | 2.041 |
|  | Past Session Information | 0.000 | 0.103 | 0.107 | 0.800 | -1.833 | 3.069 |
|  | Presenting Complaints (Symptoms) | 0.016 | 0.185 | 0.150 | 0.853 | -0.979 | 2.358 |
|  | History | 0.010 | 0.135 | 0.098 | 0.816 | -1.411 | 2.557 |
|  | Crisis Markers | 0.000 | 0.176 | 0.260 | 0.826 | -1.064 | 2.372 |
|  | Current Mental Status Examination | 0.022 | 0.335 | 0.250 | 0.848 | -0.860 | 2.803 |
|  | Psychotherapy Type | 0.145 | 0.407 | 0.369 | 0.861 | -0.883 | 2.364 |
|  | Psychotherapy Technique | 0.000 | 0.239 | 0.382 | 0.884 | -0.319 | 1.829 |
|  | Assessments | 0.000 | 0.424 | 0.841 | 0.971 | 0.603 | 0.370 |
|  | Issues Discussed in Current Session | 0.029 | 0.158 | 0.158 | 0.863 | -0.730 | 2.416 |
|  | Reflections by Therapist | 0.000 | 0.439 | 0.878 | 0.976 | 0.694 | 0.347 |
|  | Clinical Diagnosis by Reviewer | 0.000 | 0.295 | 0.549 | 0.929 | 0.073 | 1.610 |
|  | Action Plan | 0.000 | 0.276 | 0.482 | 0.911 | -0.275 | 1.246 |
|  | Next Session Details | 0.000 | 0.109 | 0.130 | 0.805 | -1.719 | 2.717 |
| **Gemma** | Patient Particulars | 0.000 | 0.144 | 0.127 | 0.813 | -1.436 | 3.006 |
|  | Clinical Identifiers | 0.000 | 0.008 | 0.003 | 0.786 | -1.828 | 2.937 |
|  | Referral Information | 0.000 | 0.035 | 0.027 | 0.802 | -1.849 | 2.781 |
|  | Therapist Information | 0.002 | 0.048 | 0.028 | 0.791 | -1.641 | 2.756 |
|  | Past Session Information | 0.000 | 0.057 | 0.047 | 0.794 | -1.898 | 3.028 |
|  | Presenting Complaints (Symptoms) | 0.009 | 0.224 | 0.132 | 0.849 | -1.046 | 2.547 |
|  | History | 0.009 | 0.104 | 0.082 | 0.813 | -1.405 | 2.574 |
|  | Crisis Markers | 0.000 | 0.017 | 0.011 | 0.804 | -1.746 | 2.628 |
|  | Current Mental Status Examination | 0.024 | 0.296 | 0.211 | 0.839 | -0.836 | 2.741 |
|  | Psychotherapy Type | 0.008 | 0.120 | 0.060 | 0.820 | -1.424 | 2.756 |
|  | Psychotherapy Technique | 0.000 | 0.056 | 0.042 | 0.805 | -1.511 | 2.510 |
|  | Assessments | 0.000 | 0.017 | 0.018 | 0.808 | -1.772 | 2.597 |
|  | Issues Discussed in Current Session | 0.020 | 0.158 | 0.146 | 0.858 | -0.623 | 2.454 |
|  | Reflections by Therapist | 0.000 | 0.000 | 0.000 | 0.798 | -1.600 | 2.711 |
|  | Clinical Diagnosis by Reviewer | 0.000 | 0.031 | 0.013 | 0.809 | -1.591 | 2.734 |
|  | Action Plan | 0.002 | 0.072 | 0.035 | 0.817 | -1.469 | 2.650 |
|  | Next Session Details | 0.000 | 0.033 | 0.019 | 0.801 | -1.895 | 2.831 |
| **Llama 3.1** | Patient Particulars | 0.020 | 0.184 | 0.098 | 0.823 | -1.453 | 2.419 |
|  | Clinical Identifiers | 0.000 | 0.043 | 0.025 | 0.782 | -1.787 | 2.422 |
|  | Referral Information | 0.000 | 0.039 | 0.024 | 0.778 | -1.766 | 2.569 |
|  | Therapist Information | 0.003 | 0.070 | 0.031 | 0.793 | -1.687 | 2.684 |
|  | Past Session Information | 0.000 | 0.080 | 0.050 | 0.808 | -1.845 | 2.596 |
|  | Presenting Complaints (Symptoms) | 0.005 | 0.181 | 0.095 | 0.835 | -1.002 | 2.232 |
|  | History | 0.007 | 0.118 | 0.081 | 0.818 | -1.502 | 2.479 |
|  | Crisis Markers | 0.000 | 0.021 | 0.012 | 0.790 | -1.715 | 2.690 |
|  | Current Mental Status Examination | 0.020 | 0.296 | 0.171 | 0.845 | -0.737 | 2.245 |
|  | Psychotherapy Type | 0.015 | 0.111 | 0.037 | 0.821 | -1.273 | 2.637 |
|  | Psychotherapy Technique | 0.000 | 0.052 | 0.020 | 0.797 | -1.144 | 2.563 |
|  | Assessments | 0.000 | 0.038 | 0.023 | 0.783 | -1.432 | 2.518 |
|  | Issues Discussed in Current Session | 0.016 | 0.179 | 0.134 | 0.847 | -0.665 | 2.375 |
|  | Reflections by Therapist | 0.000 | 0.001 | 0.001 | 0.795 | -1.551 | 2.718 |
|  | Clinical Diagnosis by Reviewer | 0.000 | 0.039 | 0.018 | 0.786 | -1.389 | 2.712 |
|  | Action Plan | 0.002 | 0.066 | 0.032 | 0.810 | -1.449 | 2.642 |
|  | Next Session Details | 0.000 | 0.048 | 0.023 | 0.795 | -1.888 | 2.643 |
| **Llama 3** | Patient Particulars | 0.018 | 0.195 | 0.102 | 0.824 | -1.506 | 2.326 |
|  | Clinical Identifiers | 0.000 | 0.045 | 0.025 | 0.784 | -1.853 | 2.183 |
|  | Referral Information | 0.000 | 0.052 | 0.030 | 0.785 | -1.891 | 2.496 |
|  | Therapist Information | 0.004 | 0.065 | 0.031 | 0.798 | -1.771 | 2.447 |
|  | Past Session Information | 0.000 | 0.089 | 0.048 | 0.808 | -1.828 | 2.526 |
|  | Presenting Complaints (Symptoms) | 0.008 | 0.178 | 0.099 | 0.835 | -0.932 | 2.231 |
|  | History | 0.015 | 0.120 | 0.081 | 0.814 | -1.587 | 2.502 |
|  | Crisis Markers | 0.000 | 0.039 | 0.021 | 0.790 | -1.752 | 2.492 |
|  | Current Mental Status Examination | 0.024 | 0.322 | 0.161 | 0.846 | -0.796 | 2.385 |
|  | Psychotherapy Type | 0.010 | 0.096 | 0.030 | 0.815 | -1.304 | 2.631 |
|  | Psychotherapy Technique | 0.000 | 0.053 | 0.021 | 0.800 | -1.075 | 2.544 |
|  | Assessments | 0.000 | 0.034 | 0.017 | 0.801 | -1.507 | 2.577 |
|  | Issues Discussed in Current Session | 0.018 | 0.199 | 0.148 | 0.849 | -0.657 | 2.332 |
|  | Reflections by Therapist | 0.000 | 0.022 | 0.011 | 0.805 | -1.609 | 2.600 |
|  | Clinical Diagnosis by Reviewer | 0.000 | 0.045 | 0.022 | 0.788 | -1.499 | 2.699 |
|  | Action Plan | 0.004 | 0.071 | 0.035 | 0.815 | -1.413 | 2.568 |
|  | Next Session Details | 0.000 | 0.044 | 0.022 | 0.794 | -1.835 | 2.650 |
| **Mentallama** | Patient Particulars | 0.006 | 0.084 | 0.046 | 0.805 | -1.465 | 2.791 |
|  | Clinical Identifiers | 0.000 | 0.006 | 0.003 | 0.797 | -1.671 | 2.590 |
|  | Referral Information | 0.000 | 0.002 | 0.001 | 0.805 | -1.548 | 2.654 |
|  | Therapist Information | 0.000 | 0.013 | 0.005 | 0.801 | -1.547 | 2.737 |
|  | Past Session Information | 0.000 | 0.071 | 0.029 | 0.812 | -1.406 | 2.375 |
|  | Presenting Complaints (Symptoms) | 0.006 | 0.167 | 0.084 | 0.837 | -0.961 | 2.606 |
|  | History | 0.003 | 0.056 | 0.042 | 0.816 | -1.341 | 2.565 |
|  | Crisis Markers | 0.000 | 0.012 | 0.005 | 0.797 | -1.631 | 2.652 |
|  | Current Mental Status Examination | 0.011 | 0.156 | 0.078 | 0.827 | -0.762 | 2.545 |
|  | Psychotherapy Type | 0.002 | 0.043 | 0.021 | 0.810 | -1.396 | 2.717 |
|  | Psychotherapy Technique | 0.000 | 0.046 | 0.016 | 0.799 | -1.283 | 2.578 |
|  | Assessments | 0.000 | 0.004 | 0.004 | 0.805 | -1.532 | 2.655 |
|  | Issues Discussed in Current Session | 0.019 | 0.139 | 0.111 | 0.856 | -0.555 | 2.432 |
|  | Reflections by Therapist | 0.000 | 0.002 | 0.001 | 0.797 | -1.579 | 2.699 |
|  | Clinical Diagnosis by Reviewer | 0.000 | 0.045 | 0.026 | 0.798 | -1.392 | 2.744 |
|  | Action Plan | 0.000 | 0.054 | 0.025 | 0.813 | -1.343 | 2.564 |
|  | Next Session Details | 0.000 | 0.012 | 0.005 | 0.805 | -1.693 | 2.594 |
| **Mistral** | Patient Particulars | 0.046 | 0.253 | 0.201 | 0.850 | -1.343 | 1.850 |
|  | Clinical Identifiers | 0.000 | 0.184 | 0.228 | 0.815 | -1.753 | 2.171 |
|  | Referral Information | 0.000 | 0.118 | 0.103 | 0.817 | -1.753 | 2.366 |
|  | Therapist Information | 0.011 | 0.180 | 0.156 | 0.820 | -1.600 | 2.316 |
|  | Past Session Information | 0.000 | 0.098 | 0.051 | 0.814 | -1.708 | 2.355 |
|  | Presenting Complaints (Symptoms) | 0.008 | 0.204 | 0.123 | 0.844 | -0.996 | 2.165 |
|  | History | 0.012 | 0.119 | 0.083 | 0.818 | -1.492 | 2.522 |
|  | Crisis Markers | 0.000 | 0.054 | 0.031 | 0.808 | -1.743 | 2.624 |
|  | Current Mental Status Examination | 0.052 | 0.372 | 0.213 | 0.870 | -0.759 | 2.059 |
|  | Psychotherapy Type | 0.026 | 0.191 | 0.085 | 0.841 | -1.285 | 2.514 |
|  | Psychotherapy Technique | 0.000 | 0.111 | 0.067 | 0.824 | -1.212 | 2.568 |
|  | Assessments | 0.000 | 0.090 | 0.066 | 0.838 | -1.711 | 2.545 |
|  | Issues Discussed in Current Session | 0.013 | 0.126 | 0.123 | 0.854 | -0.733 | 2.348 |
|  | Reflections by Therapist | 0.000 | 0.000 | 0.000 | 0.795 | -1.605 | 2.677 |
|  | Clinical Diagnosis by Reviewer | 0.000 | 0.147 | 0.225 | 0.857 | -0.765 | 2.296 |
|  | Action Plan | 0.003 | 0.082 | 0.039 | 0.816 | -1.497 | 2.640 |
|  | Next Session Details | 0.000 | 0.055 | 0.029 | 0.802 | -1.967 | 2.416 |
| **Orca 2** | Patient Particulars | 0.017 | 0.171 | 0.104 | 0.805 | -1.361 | 2.378 |
|  | Clinical Identifiers | 0.000 | 0.009 | 0.005 | 0.781 | -1.678 | 2.271 |
|  | Referral Information | 0.000 | 0.026 | 0.049 | 0.790 | -1.647 | 2.353 |
|  | Therapist Information | 0.006 | 0.115 | 0.165 | 0.818 | -1.296 | 2.168 |
|  | Past Session Information | 0.000 | 0.068 | 0.041 | 0.810 | -1.633 | 2.304 |
|  | Presenting Complaints (Symptoms) | 0.010 | 0.202 | 0.148 | 0.849 | -0.948 | 2.202 |
|  | History | 0.006 | 0.105 | 0.133 | 0.826 | -1.276 | 2.425 |
|  | Crisis Markers | 0.000 | 0.049 | 0.077 | 0.788 | -1.440 | 2.482 |
|  | Current Mental Status Examination | 0.028 | 0.293 | 0.182 | 0.825 | -0.748 | 2.143 |
|  | Psychotherapy Type | 0.024 | 0.162 | 0.142 | 0.821 | -1.076 | 2.267 |
|  | Psychotherapy Technique | 0.000 | 0.046 | 0.022 | 0.801 | -1.272 | 2.570 |
|  | Assessments | 0.000 | 0.249 | 0.493 | 0.906 | -0.303 | 1.399 |
|  | Issues Discussed in Current Session | 0.014 | 0.105 | 0.113 | 0.853 | -0.757 | 2.338 |
|  | Reflections by Therapist | 0.000 | 0.012 | 0.024 | 0.805 | -1.620 | 2.634 |
|  | Clinical Diagnosis by Reviewer | 0.000 | 0.125 | 0.196 | 0.849 | -0.777 | 2.146 |
|  | Action Plan | 0.003 | 0.081 | 0.038 | 0.818 | -1.422 | 2.620 |
|  | Next Session Details | 0.000 | 0.019 | 0.007 | 0.800 | -1.714 | 2.670 |
| **Phi 3** | Patient Particulars | 0.032 | 0.216 | 0.143 | 0.836 | -1.339 | 1.894 |
|  | Clinical Identifiers | 0.000 | 0.037 | 0.021 | 0.788 | -1.935 | 2.384 |
|  | Referral Information | 0.000 | 0.041 | 0.026 | 0.783 | -1.822 | 2.489 |
|  | Therapist Information | 0.004 | 0.070 | 0.051 | 0.799 | -1.632 | 2.438 |
|  | Past Session Information | 0.000 | 0.063 | 0.042 | 0.810 | -1.795 | 2.204 |
|  | Presenting Complaints (Symptoms) | 0.008 | 0.188 | 0.110 | 0.834 | -1.028 | 2.285 |
|  | History | 0.012 | 0.115 | 0.083 | 0.817 | -1.460 | 2.489 |
|  | Crisis Markers | 0.000 | 0.048 | 0.024 | 0.793 | -1.596 | 2.591 |
|  | Current Mental Status Examination | 0.031 | 0.314 | 0.149 | 0.850 | -0.824 | 2.064 |
|  | Psychotherapy Type | 0.019 | 0.193 | 0.115 | 0.827 | -1.356 | 2.458 |
|  | Psychotherapy Technique | 0.000 | 0.060 | 0.031 | 0.805 | -1.454 | 2.629 |
|  | Assessments | 0.000 | 0.016 | 0.009 | 0.798 | -1.707 | 2.673 |
|  | Issues Discussed in Current Session | 0.028 | 0.203 | 0.144 | 0.851 | -0.705 | 2.390 |
|  | Reflections by Therapist | 0.000 | 0.000 | 0.000 | 0.798 | -1.533 | 2.631 |
|  | Clinical Diagnosis by Reviewer | 0.000 | 0.071 | 0.078 | 0.812 | -1.299 | 2.750 |
|  | Action Plan | 0.000 | 0.067 | 0.036 | 0.815 | -1.427 | 2.568 |
|  | Next Session Details | 0.000 | 0.063 | 0.037 | 0.793 | -1.938 | 2.444 |
| **Vicuna** | Patient Particulars | 0.022 | 0.189 | 0.136 | 0.819 | -1.309 | 2.211 |
|  | Clinical Identifiers | 0.000 | 0.018 | 0.013 | 0.801 | -1.591 | 2.223 |
|  | Referral Information | 0.000 | 0.051 | 0.081 | 0.813 | -1.568 | 2.325 |
|  | Therapist Information | 0.005 | 0.070 | 0.061 | 0.807 | -1.589 | 2.458 |
|  | Past Session Information | 0.000 | 0.077 | 0.064 | 0.821 | -1.639 | 2.257 |
|  | Presenting Complaints (Symptoms) | 0.010 | 0.165 | 0.115 | 0.838 | -1.032 | 2.262 |
|  | History | 0.005 | 0.070 | 0.046 | 0.813 | -1.419 | 2.536 |
|  | Crisis Markers | 0.000 | 0.070 | 0.124 | 0.828 | -1.221 | 2.314 |
|  | Current Mental Status Examination | 0.025 | 0.246 | 0.155 | 0.845 | -0.730 | 2.212 |
|  | Psychotherapy Type | 0.013 | 0.119 | 0.091 | 0.830 | -1.162 | 2.463 |
|  | Psychotherapy Technique | 0.000 | 0.051 | 0.015 | 0.799 | -1.225 | 2.590 |
|  | Assessments | 0.000 | 0.064 | 0.122 | 0.829 | -1.235 | 2.305 |
|  | Issues Discussed in Current Session | 0.022 | 0.167 | 0.144 | 0.856 | -0.603 | 2.380 |
|  | Reflections by Therapist | 0.000 | 0.024 | 0.049 | 0.813 | -1.394 | 2.568 |
|  | Clinical Diagnosis by Reviewer | 0.000 | 0.052 | 0.028 | 0.798 | -1.413 | 2.659 |
|  | Action Plan | 0.000 | 0.042 | 0.021 | 0.806 | -1.449 | 2.626 |
|  | Next Session Details | 0.000 | 0.025 | 0.028 | 0.803 | -1.650 | 2.597 |
| **Zephyr** | Patient Particulars | 0.039 | 0.220 | 0.164 | 0.774 | -1.364 | 1.953 |
|  | Clinical Identifiers | 0.000 | 0.067 | 0.055 | 0.657 | -1.620 | 1.914 |
|  | Referral Information | 0.000 | 0.052 | 0.036 | 0.740 | -1.743 | 2.382 |
|  | Therapist Information | 0.003 | 0.061 | 0.029 | 0.679 | -1.584 | 2.120 |
|  | Past Session Information | 0.000 | 0.101 | 0.079 | 0.802 | -1.844 | 2.499 |
|  | Presenting Complaints (Symptoms) | 0.009 | 0.191 | 0.104 | 0.837 | -1.107 | 2.248 |
|  | History | 0.000 | 0.100 | 0.072 | 0.809 | -1.677 | 2.544 |
|  | Crisis Markers | 0.000 | 0.022 | 0.011 | 0.795 | -1.728 | 2.633 |
|  | Current Mental Status Examination | 0.030 | 0.267 | 0.172 | 0.783 | -0.888 | 2.022 |
|  | Psychotherapy Type | 0.028 | 0.226 | 0.202 | 0.846 | -1.037 | 2.455 |
|  | Psychotherapy Technique | 0.000 | 0.049 | 0.019 | 0.793 | -1.404 | 2.584 |
|  | Assessments | 0.000 | 0.027 | 0.021 | 0.800 | -1.623 | 2.624 |
|  | Issues Discussed in Current Session | 0.018 | 0.171 | 0.135 | 0.853 | -0.795 | 2.397 |
|  | Reflections by Therapist | 0.000 | 0.000 | 0.000 | 0.798 | -1.535 | 2.654 |
|  | Clinical Diagnosis by Reviewer | 0.000 | 0.055 | 0.042 | 0.798 | -1.388 | 2.679 |
|  | Action Plan | 0.000 | 0.065 | 0.024 | 0.811 | -1.483 | 2.584 |
|  | Next Session Details | 0.000 | 0.019 | 0.007 | 0.758 | -1.688 | 2.454 |

### **S1.8. One-shot results across different aspects of psychotherapy note summarization.**

Detailed comparison of model performance when provided with a single example for each aspect of psychotherapy note summarization. The table presents scores across six evaluation metrics (BLEU, METEOR, ROUGE-L, BERTScore, BLUERT, and InfoLM) for all 11 models on 17 aspects. Compared to zero-shot results (section S1.7), most models show significant improvements, particularly in structured aspects.

| **Model(s)** | **Aspect(s)** | **BLEU** | **METEOR** | **ROUGE-L** | **BERT** | **BLEU-RT** | **InfoLM** |
| --- | --- | --- | --- | --- | --- | --- | --- |
| **GPT4o mini** | Patient Particulars | 0.050 | 0.268 | 0.238 | 0.851 | -1.202 | 2.017 |
|  | Clinical Identifiers | 0.000 | 0.349 | 0.658 | 0.926 | -0.011 | 0.758 |
|  | Referral Information | 0.000 | 0.375 | 0.720 | 0.941 | 0.172 | 0.842 |
|  | Therapist Information | 0.041 | 0.345 | 0.569 | 0.915 | -0.221 | 1.758 |
|  | Past Session Information | 0.000 | 0.125 | 0.129 | 0.804 | -1.718 | 3.004 |
|  | Presenting Complaints (Symptoms) | 0.013 | 0.238 | 0.140 | 0.835 | -0.932 | 2.297 |
|  | History | 0.008 | 0.120 | 0.077 | 0.819 | -1.475 | 2.518 |
|  | Crisis Markers | 0.000 | 0.253 | 0.461 | 0.881 | -0.511 | 1.750 |
|  | Current Mental Status Examination | 0.049 | 0.356 | 0.228 | 0.863 | -0.712 | 2.152 |
|  | Psychotherapy Type | 0.225 | 0.358 | 0.352 | 0.874 | -0.630 | 2.165 |
|  | Psychotherapy Technique | 0.000 | 0.153 | 0.157 | 0.838 | -0.887 | 2.467 |
|  | Assessments | 0.000 | 0.398 | 0.791 | 0.966 | 0.540 | 0.731 |
|  | Issues Discussed in Current Session | 0.020 | 0.215 | 0.160 | 0.853 | -0.547 | 2.350 |
|  | Reflections by the Therapist | 0.000 | 0.341 | 0.683 | 0.935 | 0.245 | 0.871 |
|  | Clinical Diagnosis by Reviewer | 0.000 | 0.169 | 0.250 | 0.856 | -0.655 | 2.222 |
|  | Action Plan | 0.000 | 0.255 | 0.412 | 0.893 | -0.408 | 1.397 |
|  | Next Session Details | 0.000 | 0.120 | 0.182 | 0.804 | -1.705 | 2.805 |
| **Gemini Pro** | Patient Particulars | 0.035 | 0.252 | 0.280 | 0.856 | -1.014 | 2.265 |
|  | Clinical Identifiers | 0.000 | 0.274 | 0.442 | 0.868 | -0.747 | 1.575 |
|  | Referral Information | 0.000 | 0.381 | 0.692 | 0.915 | 0.054 | 1.048 |
|  | Therapist Information | 0.016 | 0.327 | 0.538 | 0.902 | -0.225 | 1.697 |
|  | Past Session Information | 0.000 | 0.115 | 0.149 | 0.812 | -1.657 | 2.837 |
|  | Presenting Complaints (Symptoms) | 0.010 | 0.206 | 0.170 | 0.857 | -0.951 | 2.332 |
|  | History | 0.007 | 0.145 | 0.140 | 0.831 | -1.224 | 2.429 |
|  | Crisis Markers | 0.000 | 0.387 | 0.733 | 0.912 | 0.298 | 0.949 |
|  | Current Mental Status Examination | 0.040 | 0.337 | 0.269 | 0.859 | -0.771 | 2.482 |
|  | Psychotherapy Type | 0.205 | 0.414 | 0.426 | 0.874 | -0.670 | 2.014 |
|  | Psychotherapy Technique | 0.000 | 0.174 | 0.318 | 0.869 | -0.467 | 1.909 |
|  | Assessments | 0.000 | 0.400 | 0.789 | 0.940 | 0.485 | 0.752 |
|  | Issues Discussed in Current Session | 0.021 | 0.188 | 0.156 | 0.856 | -0.648 | 2.456 |
|  | Reflections by the Therapist | 0.000 | 0.252 | 0.504 | 0.881 | -0.272 | 1.445 |
|  | Clinical Diagnosis by Reviewer | 0.000 | 0.275 | 0.496 | 0.926 | -0.059 | 1.653 |
|  | Action Plan | 0.000 | 0.254 | 0.439 | 0.899 | -0.408 | 1.714 |
|  | Next Session Details | 0.000 | 0.266 | 0.497 | 0.890 | -0.505 | 1.372 |
| **Gemma** | Patient Particulars | 0.003 | 0.054 | 0.037 | 0.803 | -1.421 | 2.751 |
|  | Clinical Identifiers | 0.000 | 0.007 | 0.002 | 0.796 | -1.658 | 2.716 |
|  | Referral Information | 0.000 | 0.004 | 0.002 | 0.805 | -1.633 | 2.696 |
|  | Therapist Information | 0.000 | 0.008 | 0.003 | 0.800 | -1.659 | 2.748 |
|  | Past Session Information | 0.000 | 0.047 | 0.022 | 0.803 | -1.560 | 2.452 |
|  | Presenting Complaints (Symptoms) | 0.009 | 0.160 | 0.111 | 0.850 | -0.888 | 2.421 |
|  | History | 0.000 | 0.078 | 0.067 | 0.824 | -1.198 | 2.518 |
|  | Crisis Markers | 0.000 | 0.010 | 0.014 | 0.805 | -1.645 | 2.666 |
|  | Current Mental Status Examination | 0.004 | 0.085 | 0.062 | 0.825 | -0.880 | 2.502 |
|  | Psychotherapy Type | 0.000 | 0.018 | 0.008 | 0.807 | -1.474 | 2.698 |
|  | Psychotherapy Technique | 0.000 | 0.029 | 0.015 | 0.806 | -1.484 | 2.520 |
|  | Assessments | 0.000 | 0.005 | 0.011 | 0.806 | -1.697 | 2.657 |
|  | Issues Discussed in Current Session | 0.035 | 0.198 | 0.168 | 0.866 | -0.445 | 2.390 |
|  | Reflections by the Therapist | 0.000 | 0.000 | 0.002 | 0.801 | -1.625 | 2.659 |
|  | Clinical Diagnosis by Reviewer | 0.000 | 0.026 | 0.014 | 0.803 | -1.466 | 2.753 |
|  | Action Plan | 0.000 | 0.085 | 0.041 | 0.819 | -1.406 | 2.667 |
|  | Next Session Details | 0.000 | 0.016 | 0.005 | 0.802 | -1.596 | 2.604 |
| **Llama 3.1** | Patient Particulars | 0.031 | 0.206 | 0.134 | 0.832 | -1.450 | 1.998 |
|  | Clinical Identifiers | 0.000 | 0.050 | 0.031 | 0.786 | -2.005 | 2.252 |
|  | Referral Information | 0.000 | 0.047 | 0.030 | 0.790 | -2.047 | 2.575 |
|  | Therapist Information | 0.003 | 0.066 | 0.039 | 0.789 | -1.881 | 2.518 |
|  | Past Session Information | 0.000 | 0.072 | 0.046 | 0.813 | -1.836 | 2.376 |
|  | Presenting Complaints (Symptoms) | 0.013 | 0.196 | 0.107 | 0.842 | -0.959 | 2.272 |
|  | History | 0.013 | 0.096 | 0.073 | 0.816 | -1.496 | 2.535 |
|  | Crisis Markers | 0.000 | 0.042 | 0.024 | 0.796 | -2.197 | 2.725 |
|  | Current Mental Status Examination | 0.041 | 0.308 | 0.174 | 0.851 | -0.903 | 1.988 |
|  | Psychotherapy Type | 0.015 | 0.119 | 0.035 | 0.808 | -1.988 | 2.667 |
|  | Psychotherapy Technique | 0.000 | 0.047 | 0.022 | 0.793 | -1.748 | 2.585 |
|  | Assessments | 0.000 | 0.061 | 0.048 | 0.791 | -1.802 | 2.569 |
|  | Issues Discussed in Current Session | 0.027 | 0.205 | 0.150 | 0.854 | -0.595 | 2.394 |
|  | Reflections by the Therapist | 0.000 | 0.012 | 0.007 | 0.795 | -1.749 | 2.670 |
|  | Clinical Diagnosis by Reviewer | 0.000 | 0.035 | 0.020 | 0.783 | -1.754 | 2.641 |
|  | Action Plan | 0.000 | 0.054 | 0.032 | 0.812 | -1.447 | 2.551 |
|  | Next Session Details | 0.000 | 0.031 | 0.017 | 0.794 | -1.909 | 2.599 |
| **Llama 3** | Patient Particulars | 0.026 | 0.190 | 0.104 | 0.830 | -1.510 | 2.182 |
|  | Clinical Identifiers | 0.000 | 0.051 | 0.031 | 0.787 | -2.021 | 2.177 |
|  | Referral Information | 0.000 | 0.052 | 0.031 | 0.786 | -1.996 | 2.478 |
|  | Therapist Information | 0.004 | 0.064 | 0.034 | 0.794 | -1.869 | 2.383 |
|  | Past Session Information | 0.000 | 0.069 | 0.043 | 0.815 | -1.723 | 2.333 |
|  | Presenting Complaints (Symptoms) | 0.013 | 0.207 | 0.104 | 0.841 | -1.005 | 2.201 |
|  | History | 0.013 | 0.095 | 0.069 | 0.812 | -1.477 | 2.493 |
|  | Crisis Markers | 0.000 | 0.048 | 0.027 | 0.794 | -1.919 | 2.615 |
|  | Current Mental Status Examination | 0.045 | 0.326 | 0.166 | 0.856 | -0.721 | 1.997 |
|  | Psychotherapy Type | 0.015 | 0.123 | 0.033 | 0.812 | -1.654 | 2.497 |
|  | Psychotherapy Technique | 0.000 | 0.048 | 0.023 | 0.796 | -1.547 | 2.567 |
|  | Assessments | 0.000 | 0.042 | 0.025 | 0.793 | -2.171 | 2.687 |
|  | Issues Discussed in Current Session | 0.022 | 0.213 | 0.148 | 0.853 | -0.651 | 2.386 |
|  | Reflections by the Therapist | 0.000 | 0.009 | 0.005 | 0.798 | -1.584 | 2.666 |
|  | Clinical Diagnosis by Reviewer | 0.000 | 0.026 | 0.016 | 0.787 | -1.660 | 2.615 |
|  | Action Plan | 0.000 | 0.052 | 0.032 | 0.811 | -1.434 | 2.533 |
|  | Next Session Details | 0.000 | 0.036 | 0.017 | 0.799 | -1.835 | 2.525 |
| **Mentallama** | Patient Particulars | 0.014 | 0.096 | 0.119 | 0.804 | -1.280 | 2.640 |
|  | Clinical Identifiers | 0.000 | 0.000 | 0.000 | 0.810 | -1.471 | 2.398 |
|  | Referral Information | 0.000 | 0.024 | 0.049 | 0.812 | -1.326 | 2.355 |
|  | Therapist Information | 0.000 | 0.009 | 0.000 | 0.799 | -1.470 | 2.544 |
|  | Past Session Information | 0.000 | 0.032 | 0.031 | 0.794 | -1.451 | 2.334 |
|  | Presenting Complaints (Symptoms) | 0.017 | 0.110 | 0.072 | 0.777 | -1.192 | 2.461 |
|  | History | 0.002 | 0.035 | 0.047 | 0.794 | -1.398 | 2.618 |
|  | Crisis Markers | 0.000 | 0.004 | 0.001 | 0.790 | -1.513 | 2.608 |
|  | Current Mental Status Examination | 0.021 | 0.103 | 0.084 | 0.803 | -1.115 | 2.445 |
|  | Psychotherapy Type | 0.005 | 0.045 | 0.036 | 0.807 | -1.282 | 2.511 |
|  | Psychotherapy Technique | 0.000 | 0.011 | 0.002 | 0.790 | -1.421 | 2.501 |
|  | Assessments | 0.000 | 0.003 | 0.002 | 0.793 | -1.493 | 2.550 |
|  | Issues Discussed in Current Session | 0.018 | 0.096 | 0.079 | 0.801 | -1.060 | 2.549 |
|  | Reflections by the Therapist | 0.000 | 0.000 | 0.000 | 0.795 | -1.453 | 2.635 |
|  | Clinical Diagnosis by Reviewer | 0.000 | 0.008 | 0.013 | 0.787 | -1.377 | 2.577 |
|  | Action Plan | 0.002 | 0.040 | 0.026 | 0.778 | -1.432 | 2.520 |
|  | Next Session Details | 0.000 | 0.036 | 0.054 | 0.824 | -1.304 | 2.384 |
| **Mistral** | Patient Particulars | 0.049 | 0.253 | 0.205 | 0.851 | -1.356 | 1.950 |
|  | Clinical Identifiers | 0.000 | 0.146 | 0.158 | 0.811 | -1.767 | 2.204 |
|  | Referral Information | 0.000 | 0.113 | 0.090 | 0.822 | -1.840 | 2.462 |
|  | Therapist Information | 0.010 | 0.165 | 0.142 | 0.820 | -1.528 | 2.348 |
|  | Past Session Information | 0.000 | 0.129 | 0.067 | 0.833 | -1.682 | 2.309 |
|  | Presenting Complaints (Symptoms) | 0.015 | 0.208 | 0.152 | 0.851 | -0.886 | 2.252 |
|  | History | 0.012 | 0.105 | 0.078 | 0.820 | -1.437 | 2.511 |
|  | Crisis Markers | 0.000 | 0.049 | 0.038 | 0.807 | -1.773 | 2.626 |
|  | Current Mental Status Examination | 0.048 | 0.305 | 0.192 | 0.863 | -0.739 | 2.121 |
|  | Psychotherapy Type | 0.033 | 0.230 | 0.167 | 0.851 | -1.095 | 2.532 |
|  | Psychotherapy Technique | 0.000 | 0.110 | 0.104 | 0.832 | -1.091 | 2.568 |
|  | Assessments | 0.000 | 0.110 | 0.106 | 0.832 | -1.567 | 2.507 |
|  | Issues Discussed in Current Session | 0.036 | 0.178 | 0.156 | 0.863 | -0.630 | 2.349 |
|  | Reflections by the Therapist | 0.000 | 0.000 | 0.000 | 0.798 | -1.605 | 2.695 |
|  | Clinical Diagnosis by Reviewer | 0.000 | 0.138 | 0.211 | 0.856 | -0.745 | 2.333 |
|  | Action Plan | 0.003 | 0.073 | 0.034 | 0.815 | -1.478 | 2.574 |
|  | Next Session Details | 0.000 | 0.073 | 0.047 | 0.808 | -1.913 | 2.381 |
| **Orca 2** | Patient Particulars | 0.000 | 0.073 | 0.090 | 0.758 | -1.335 | 2.398 |
|  | Clinical Identifiers | 0.000 | 0.122 | 0.244 | 0.775 | -0.931 | 1.623 |
|  | Referral Information | 0.000 | 0.160 | 0.318 | 0.799 | -0.690 | 1.519 |
|  | Therapist Information | 0.000 | 0.054 | 0.100 | 0.755 | -1.221 | 2.086 |
|  | Past Session Information | 0.000 | 0.078 | 0.114 | 0.776 | -1.247 | 2.040 |
|  | Presenting Complaints (Symptoms) | 0.000 | 0.102 | 0.086 | 0.790 | -1.217 | 2.271 |
|  | History | 0.000 | 0.027 | 0.029 | 0.776 | -1.523 | 2.533 |
|  | Crisis Markers | 0.000 | 0.160 | 0.317 | 0.841 | -0.672 | 1.782 |
|  | Current Mental Status Examination | 0.039 | 0.107 | 0.105 | 0.760 | -1.221 | 2.515 |
|  | Psychotherapy Type | 0.007 | 0.094 | 0.176 | 0.795 | -1.006 | 2.092 |
|  | Psychotherapy Technique | 0.000 | 0.082 | 0.132 | 0.746 | -1.068 | 2.135 |
|  | Assessments | 0.000 | 0.165 | 0.317 | 0.850 | -0.637 | 1.776 |
|  | Issues Discussed in Current Session | 0.012 | 0.087 | 0.067 | 0.742 | -1.288 | 2.249 |
|  | Reflections by the Therapist | 0.000 | 0.110 | 0.220 | 0.789 | -0.824 | 1.624 |
|  | Clinical Diagnosis by Reviewer | 0.000 | 0.073 | 0.146 | 0.836 | -0.945 | 2.037 |
|  | Action Plan | 0.000 | 0.062 | 0.083 | 0.785 | -1.326 | 2.186 |
|  | Next Session Details | 0.000 | 0.079 | 0.149 | 0.813 | -1.210 | 2.115 |
| **Phi 3** | Patient Particulars | 0.028 | 0.224 | 0.170 | 0.836 | -1.315 | 2.127 |
|  | Clinical Identifiers | 0.000 | 0.042 | 0.044 | 0.794 | -1.823 | 2.395 |
|  | Referral Information | 0.000 | 0.042 | 0.042 | 0.797 | -1.789 | 2.484 |
|  | Therapist Information | 0.000 | 0.088 | 0.081 | 0.803 | -1.468 | 2.394 |
|  | Past Session Information | 0.000 | 0.090 | 0.066 | 0.824 | -1.611 | 2.377 |
|  | Presenting Complaints (Symptoms) | 0.007 | 0.181 | 0.098 | 0.836 | -0.950 | 2.351 |
|  | History | 0.013 | 0.103 | 0.068 | 0.816 | -1.341 | 2.432 |
|  | Crisis Markers | 0.000 | 0.090 | 0.085 | 0.800 | -1.508 | 2.533 |
|  | Current Mental Status Examination | 0.056 | 0.329 | 0.183 | 0.852 | -0.814 | 2.046 |
|  | Psychotherapy Type | 0.040 | 0.257 | 0.238 | 0.851 | -1.135 | 2.485 |
|  | Psychotherapy Technique | 0.000 | 0.120 | 0.124 | 0.823 | -1.143 | 2.512 |
|  | Assessments | 0.000 | 0.023 | 0.037 | 0.826 | -1.440 | 2.426 |
|  | Issues Discussed in Current Session | 0.029 | 0.215 | 0.161 | 0.859 | -0.608 | 2.400 |
|  | Reflections by the Therapist | 0.000 | 0.028 | 0.049 | 0.807 | -1.494 | 2.519 |
|  | Clinical Diagnosis by Reviewer | 0.000 | 0.082 | 0.122 | 0.825 | -1.247 | 2.681 |
|  | Action Plan | 0.002 | 0.067 | 0.034 | 0.813 | -1.388 | 2.528 |
|  | Next Session Details | 0.000 | 0.033 | 0.014 | 0.806 | -1.849 | 2.600 |
| **Vicuna** | Patient Particulars | 0.008 | 0.084 | 0.109 | 0.790 | -1.293 | 2.533 |
|  | Clinical Identifiers | 0.000 | 0.030 | 0.052 | 0.778 | -1.388 | 2.299 |
|  | Referral Information | 0.000 | 0.012 | 0.024 | 0.787 | -1.431 | 2.491 |
|  | Therapist Information | 0.000 | 0.011 | 0.004 | 0.793 | -1.394 | 2.579 |
|  | Past Session Information | 0.000 | 0.035 | 0.074 | 0.797 | -1.261 | 2.391 |
|  | Presenting Complaints (Symptoms) | 0.005 | 0.099 | 0.061 | 0.788 | -1.241 | 2.323 |
|  | History | 0.004 | 0.030 | 0.021 | 0.766 | -1.425 | 2.631 |
|  | Crisis Markers | 0.000 | 0.028 | 0.050 | 0.793 | -1.341 | 2.429 |
|  | Current Mental Status Examination | 0.033 | 0.119 | 0.095 | 0.797 | -1.179 | 2.496 |
|  | Psychotherapy Type | 0.003 | 0.048 | 0.035 | 0.802 | -1.313 | 2.542 |
|  | Psychotherapy Technique | 0.000 | 0.043 | 0.024 | 0.796 | -1.216 | 2.424 |
|  | Assessments | 0.000 | 0.028 | 0.050 | 0.795 | -1.327 | 2.548 |
|  | Issues Discussed in Current Session | 0.016 | 0.092 | 0.069 | 0.787 | -1.139 | 2.546 |
|  | Reflections by the Therapist | 0.000 | 0.012 | 0.024 | 0.794 | -1.377 | 2.631 |
|  | Clinical Diagnosis by Reviewer | 0.000 | 0.004 | 0.002 | 0.773 | -1.418 | 2.520 |
|  | Action Plan | 0.000 | 0.037 | 0.035 | 0.790 | -1.394 | 2.613 |
|  | Next Session Details | 0.000 | 0.022 | 0.030 | 0.793 | -1.473 | 2.504 |
| **Zephyr** | Patient Particulars | 0.014 | 0.124 | 0.076 | 0.805 | -1.527 | 2.400 |
|  | Clinical Identifiers | 0.000 | 0.025 | 0.020 | 0.767 | -1.895 | 2.413 |
|  | Referral Information | 0.000 | 0.026 | 0.021 | 0.771 | -1.881 | 2.535 |
|  | Therapist Information | 0.000 | 0.026 | 0.013 | 0.766 | -1.854 | 2.522 |
|  | Past Session Information | 0.000 | 0.056 | 0.036 | 0.792 | -1.861 | 2.520 |
|  | Presenting Complaints (Symptoms) | 0.008 | 0.132 | 0.069 | 0.818 | -1.231 | 2.436 |
|  | History | 0.010 | 0.064 | 0.048 | 0.784 | -1.529 | 2.507 |
|  | Crisis Markers | 0.000 | 0.023 | 0.016 | 0.791 | -1.827 | 2.577 |
|  | Current Mental Status Examination | 0.035 | 0.222 | 0.147 | 0.826 | -1.044 | 2.117 |
|  | Psychotherapy Type | 0.009 | 0.097 | 0.063 | 0.800 | -1.624 | 2.590 |
|  | Psychotherapy Technique | 0.000 | 0.026 | 0.008 | 0.784 | -1.643 | 2.567 |
|  | Assessments | 0.000 | 0.030 | 0.027 | 0.794 | -1.876 | 2.658 |
|  | Issues Discussed in Current Session | 0.022 | 0.149 | 0.115 | 0.836 | -0.885 | 2.450 |
|  | Reflections by the Therapist | 0.000 | 0.022 | 0.020 | 0.772 | -1.739 | 2.627 |
|  | Clinical Diagnosis by Reviewer | 0.000 | 0.015 | 0.013 | 0.784 | -1.606 | 2.712 |
|  | Action Plan | 0.000 | 0.032 | 0.021 | 0.795 | -1.637 | 2.618 |
|  | Next Session Details | 0.000 | 0.015 | 0.010 | 0.789 | -1.827 | 2.669 |

### **S1.9. Statistical comparison of expert (TRACE) ratings across the three best-performing LLMs and the human gold standard.**

Ratings were pooled across six expert evaluators over 168 sessions (n=301 rating instances per system per domain). The primary analysis is the Friedman test for matched samples (n=296 complete quadruples) with Nemenyi post-hoc comparisons (Panels A and B); the Kruskal–Wallis analysis, which treats the systems as independent, is retained as a sensitivity analysis (Panels E and F). Omnibus p-values are reported both raw and Holm–Bonferroni-corrected across the five TRACE domains; the pooled overall score is reported separately and is not included in that correction family. Nemenyi post-hoc p-values are family-wise corrected within each domain. Significant comparisons (p<0.05) are those to be interpreted; α is ordinal Krippendorff's α with bootstrap 95% CI (1000 resamples).

**Panel A. Friedman omnibus tests, primary analysis (df=3, four matched systems: GPT-4o-mini, Gemini Pro, Mistral, Human; n=296 complete quadruples)**

| **Domain** | **H** | **df** | **p (raw)** | **p (Holm)** |
| --- | --- | --- | --- | --- |
| Trustworthiness | 17.00 | 3 | 7.1×10⁻⁴ | 0.001 |
| Relevance | 61.64 | 3 | 2.6×10⁻¹³ | 1.3×10⁻¹² |
| Accuracy | 39.69 | 3 | 1.2×10⁻⁸ | 5.0×10⁻⁸ |
| Comprehensiveness | 2.79 | 3 | 0.424 | 0.424 |
| Expression | 35.86 | 3 | 8.0×10⁻⁸ | 2.4×10⁻⁷ |
| Overall (pooled) | 31.18 | 3 | 7.8×10⁻⁷ | – |

**Panel B. Nemenyi post-hoc pairwise p-values, primary analysis (following the Friedman test, n=296)**

**Trustworthiness**

| **Comparison** | **p** |
| --- | --- |
| GPT-4o mini vs Gemini Pro | 0.988 |
| GPT-4o mini vs Mistral | 0.407 |
| GPT-4o mini vs Human | 0.003 |
| Gemini Pro vs Mistral | 0.616 |
| Gemini Pro vs Human | 0.010 |
| Mistral vs Human | 0.255 |

**Relevance**

| **Comparison** | **p** |
| --- | --- |
| GPT-4o mini vs Gemini Pro | 0.827 |
| GPT-4o mini vs Mistral | 0.333 |
| GPT-4o mini vs Human | <0.001 |
| Gemini Pro vs Mistral | 0.847 |
| Gemini Pro vs Human | <0.001 |
| Mistral vs Human | <0.001 |

**Accuracy**

| **Comparison** | **p** |
| --- | --- |
| GPT-4o mini vs Gemini Pro | 0.696 |
| GPT-4o mini vs Mistral | 0.074 |
| GPT-4o mini vs Human | <0.001 |
| Gemini Pro vs Mistral | 0.002 |
| Gemini Pro vs Human | <0.001 |
| Mistral vs Human | 0.321 |

**Comprehensiveness**

| **Comparison** | **p** |
| --- | --- |
| GPT-4o mini vs Gemini Pro | 0.999 |
| GPT-4o mini vs Mistral | 0.537 |
| GPT-4o mini vs Human | 0.878 |
| Gemini Pro vs Mistral | 0.613 |
| Gemini Pro vs Human | 0.923 |
| Mistral vs Human | 0.935 |

**Expression**

| **Comparison** | **p** |
| --- | --- |
| GPT-4o mini vs Gemini Pro | 0.806 |
| GPT-4o mini vs Mistral | 0.587 |
| GPT-4o mini vs Human | <0.001 |
| Gemini Pro vs Mistral | 0.984 |
| Gemini Pro vs Human | <0.001 |
| Mistral vs Human | 0.001 |

**Overall (pooled)**

| **Comparison** | **p** |
| --- | --- |
| GPT-4o mini vs Gemini Pro | 0.988 |
| GPT-4o mini vs Mistral | 0.249 |
| GPT-4o mini vs Human | <0.001 |
| Gemini Pro vs Mistral | 0.427 |
| Gemini Pro vs Human | <0.001 |
| Mistral vs Human | 0.032 |

**Panel C. Inter-rater agreement (ordinal Krippendorff's α, 460 doubly-coded units)**

| **Domain** | **α** | **95% CI** |
| --- | --- | --- |
| Trustworthiness | 0.194 | 0.112 to 0.274 |
| Relevance | 0.084 | -0.007 to 0.169 |
| Accuracy | 0.138 | 0.055 to 0.217 |
| Comprehensiveness | 0.052 | -0.042 to 0.138 |
| Expression | 0.035 | -0.046 to 0.121 |
| Overall (pooled) | 0.100 | 0.014 to 0.176 |

**Panel D.** Descriptive statistics underlying Table 4 of the main manuscript (n=301 rating instances per system per domain). Mean (SD) reproduces the values shown in Table 4 of the main manuscript. Median and IQR describe the same ratings on the ordinal scale and are near-constant across systems, which is what Table 4 of the main manuscript reports means. Mean rank is the average rank of that system's 301 ratings among all 1,204 ratings pooled within the domain (possible range 1–1,204, midpoint 602.5; higher is better) and is the quantity the Friedman and Nemenyi tests compare.

| **Domain** | **System** | **Mean (SD)** | **Median** | **IQR** | **Mean rank** |
| --- | --- | --- | --- | --- | --- |
| Trustworthiness | Gemini Pro | 2.78 (0.83) | 3.0 | 2-3 | 573.4 |
|  | GPT-4o mini | 2.76 (0.91) | 3.0 | 2-3 | 563.7 |
|  | Mistral | 2.88 (0.88) | 3.0 | 2-4 | 609.4 |
|  | Human | 3.01 (0.97) | 3.0 | 2-4 | 663.5 |
| Relevance | Gemini Pro | 2.65 (0.94) | 3.0 | 2-3 | 560.9 |
|  | GPT-4o mini | 2.58 (0.95) | 2.0 | 2-3 | 535.4 |
|  | Mistral | 2.71 (0.86) | 3.0 | 2-3 | 585.2 |
|  | Human | 3.09 (0.88) | 3.0 | 3-4 | 728.6 |
| Accuracy | Gemini Pro | 2.60 (0.85) | 3.0 | 2-3 | 530.1 |
|  | GPT-4o mini | 2.67 (0.95) | 3.0 | 2-3 | 562.5 |
|  | Mistral | 2.88 (0.87) | 3.0 | 2-4 | 633.5 |
|  | Human | 2.99 (0.93) | 3.0 | 2-4 | 684.0 |
| Comprehensiveness | Gemini Pro | 2.80 (0.90) | 3.0 | 2-3 | 589.7 |
|  | GPT-4o mini | 2.78 (0.90) | 3.0 | 2-3 | 586.2 |
|  | Mistral | 2.88 (0.85) | 3.0 | 2-4 | 625.8 |
|  | Human | 2.83 (0.95) | 3.0 | 2-4 | 608.3 |
| Expression | Gemini Pro | 2.88 (0.97) | 3.0 | 2-4 | 575.7 |
|  | GPT-4o mini | 2.78 (0.96) | 3.0 | 2-4 | 549.0 |
|  | Mistral | 2.90 (0.87) | 3.0 | 2-4 | 586.6 |
|  | Human | 3.19 (0.90) | 3.0 | 3-4 | 698.7 |
| Overall | Gemini Pro | 2.74 (0.80) | 2.8 | 2-3 | 560.5 |
|  | GPT-4o mini | 2.72 (0.84) | 2.6 | 2-3 | 550.3 |
|  | Mistral | 2.85 (0.74) | 2.8 | 2-3 | 607.6 |
|  | Human | 3.02 (0.84) | 3.0 | 3-4 | 691.6 |

**Panel E. Kruskal–Wallis omnibus tests (sensitivity analysis; treats systems as independent, df=3, n=301 per group)**

| **Domain** | **H** | **p (raw)** | **p (Holm)** |
| --- | --- | --- | --- |
| Trustworthiness | 17.00 | 7.1×10⁻⁴ | 0.001 |
| Relevance | 61.64 | 2.6×10⁻¹³ | 1.3×10⁻¹² |
| Accuracy | 39.69 | 1.2×10⁻⁸ | 5.0×10⁻⁸ |
| Comprehensiveness | 2.79 | 0.424 | 0.424 |
| Expression | 35.86 | 8.0×10⁻⁸ | 2.4×10⁻⁷ |
| Overall (pooled) | 31.18 | 7.8×10⁻⁷ | – |

**Panel F. Nemenyi post-hoc pairwise p-values (Kruskal–Wallis sensitivity analysis)**

**Trustworthiness**

| **Comparison** | **p** |
| --- | --- |
| GPT-4o mini vs Gemini Pro | 0.988 |
| GPT-4o mini vs Mistral | 0.407 |
| GPT-4o mini vs Human | 0.003 |
| Gemini Pro vs Mistral | 0.616 |
| Gemini Pro vs Human | 0.010 |
| Mistral vs Human | 0.255 |

**Relevance**

| **Comparison** | **p** |
| --- | --- |
| GPT-4o mini vs Gemini Pro | 0.827 |
| GPT-4o mini vs Mistral | 0.333 |
| GPT-4o mini vs Human | <0.001 |
| Gemini Pro vs Mistral | 0.847 |
| Gemini Pro vs Human | <0.001 |
| Mistral vs Human | <0.001 |

**Accuracy**

| **Comparison** | **p** |
| --- | --- |
| GPT-4o mini vs Gemini Pro | 0.696 |
| GPT-4o mini vs Mistral | 0.074 |
| GPT-4o mini vs Human | <0.001 |
| Gemini Pro vs Mistral | 0.002 |
| Gemini Pro vs Human | <0.001 |
| Mistral vs Human | 0.321 |

**Comprehensiveness**

| **Comparison** | **p** |
| --- | --- |
| GPT-4o mini vs Gemini Pro | 0.999 |
| GPT-4o mini vs Mistral | 0.537 |
| GPT-4o mini vs Human | 0.878 |
| Gemini Pro vs Mistral | 0.613 |
| Gemini Pro vs Human | 0.923 |
| Mistral vs Human | 0.935 |

**Expression**

| **Comparison** | **p** |
| --- | --- |
| GPT-4o mini vs Gemini Pro | 0.806 |
| GPT-4o mini vs Mistral | 0.587 |
| GPT-4o mini vs Human | <0.001 |
| Gemini Pro vs Mistral | 0.984 |
| Gemini Pro vs Human | <0.001 |
| Mistral vs Human | 0.001 |

**Overall (pooled)**

| **Comparison** | **p** |
| --- | --- |
| GPT-4o mini vs Gemini Pro | 0.988 |
| GPT-4o mini vs Mistral | 0.249 |
| GPT-4o mini vs Human | <0.001 |
| Gemini Pro vs Mistral | 0.427 |
| Gemini Pro vs Human | <0.001 |
| Mistral vs Human | 0.032 |
