## Supplementary Document 2 for "Clinically Grounded AI-Scribing in Psychotherapy: Benchmarking LLMs Against Expert Documentation in the iCARE Framework"

Multimedia Appendix 2. AI-assisted documentation in psychotherapy: characteristics, technology and findings of prior studies

Clinically Grounded AI-Scribing in Psychotherapy: Benchmarking LLMs Against Expert Documentation in the iCARE Framework

This appendix supports the Prior Work subsection of the Introduction of the main manuscript, where it is cited as Multimedia Appendix 2. It is presented as a multimedia appendix rather than as a table in the main text because its length exceeds the limit for main-text tables. Reference numbers in the table refer to the reference list at the end of this appendix, not to the reference list of the main manuscript.

Abbreviations used in this appendix: AI (artificial intelligence); ASR (automatic speech recognition); CBT (cognitive behavioral therapy); LLM (large language model); NLP (natural language processing); RCT (randomized controlled trial).

| **Study (country)** | **Psychotherapy type/note type** | **Data source** | **Design** | **Technology/ system and LLM/NLP techniques** | **AI documentation function** | **Main**  **finding** |
| --- | --- | --- | --- | --- | --- | --- |
| **Perlich & Meinel, 2015 (Germany)**[1] | **Psychotherapy / session notes** | Therapist demonstration sessions in a psychotherapy clinic; no fixed patient dataset | Exploratory experimental study | **Tele-Board MED;**  digital whiteboard with client participation | Automatic treatment- session summary generation | Demonstrated feasibility of generating structured psychotherapy session summaries |
| **Sadeh-Sharvit et al., 2022 (USA)**[2] | **Behavioral therapy / progress or session notes** | Real-world anonymized Eleos dataset of **17,607 sessions**, **322 therapists**, **3519 clients** | Retrospective observational study | **Eleos platform**; ASR, speaker diarization, VADER-based NLP^1^ | Analysis of session- summary use in routine care | Session summaries were associated with faster note completion |
| **Srivastava et al., 2022 (India, USA)**[3] | **Counseling or psychotherapy dialogue / session notes** | **HOPE** dataset expanded to **MEMO** with 12.9K expert- annotated utterances | Comparative experimental study | **ConSum / MEMO pipeline**; BART, T5, knowledge-guided utterance filtering^2^ | Knowledge-guided counseling summarization | Domain-informed filtering improved the relevance and meaningfulness of summaries |
| **Sadeh-Sharvit et al., 2023 (USA)**[4] | **CBT / progress or session notes** | RCT of **47 adult CBT patients** with session audio, summaries, PHQ-9 and GAD-7 data | Randomized controlled trial (AI Scribing vs TaU)^3^ | **Eleos platform**; AI-assisted transcription and summarization | AI-supported summarization and progress-note drafting | Strongest direct clinical evidence; AI support improved workflow efficiency and symptom outcomes |
| **Wu et al., 2023 (Taiwan)**[5] | **EMDR teletherapy / progress or session notes^4^** | System-generated real-world and simulated teletherapy session data from **24 participants** | System development with feasibility and usability testing | AI-augmented teletherapy platform; **ChatGPT-3.5**, **Claude-3.5 Sonnet**, emotion- recognition module | AI-generated post-session summaries and reports | Reduced documentation time by >50% and supported remote psychotherapy workflow |
| **Adhikary et al., 2024 (India)**[6] | **Psychotherapy or counseling / progress or session notes** | **MEMO** dataset and **MentalCLOUDS** aspect- level counseling summaries^5^ | Benchmarking study | 11 LLMs - **BART, T5, GPT- family, Mistral, MentalLlama, MentalBART** | LLM-based aspect-wise session summarization | Domain-specific models outperformed general-purpose models for counseling summarization |
| **Srivastava et al., 2024 (India)**[7] | **Counseling / progress or session notes** | **MEMO** dataset (primary) and **ACI-BENCH^6^** | Experimental study | **PIECE framework**; BART, Pegasus, T5, Flan-T5, Mistral-7B, Zephyr-7B, Llama-2, MentalLlama | Knowledge planning-guided LLM summarization | Improved counseling note generation & reduced hallucinations |
| **Lee et al., 2025 (South Korea)**[8] | **Child counseling / session notes** | Video-recorded child counseling session data; Korean Children’s Voice Records Dataset | Mixed-method study | ASR, Child voice-aware-STT **(Whisper-large-v2),** AI-video captioning **(GPT-4o)**, Speaker recognition **(Speechbrain)** | Verbal Expression Inference, Non-Verbal Expression Inference, editable dashboard, summarization | Improved transcription workflow and was perceived as useful by counselors |
| **Keerthana & Gupta, 2025 (USA)**[9] | **No specific psychotherapy** | Longitudinal heterogeneous clinical notes across hospital visits, **MIMIC- III**-based.^7^ | Experimental study | **DENSE;** CLI-RAG framework; No specific LLM mentioned | Longitudinal SOAP style progress-note generation across visits with temporal continuity and clinically grounded synthesis | Demonstrated strong potential for longitudinal clinical documentation. |
| **Kumar et al., 2025 (Germany & India)**[10] | **Motivational interviewing / progress or session notes** | **AnnoMI** and **AnnoSUM- MI** - expert-annotated MI datasets.^8^ | Experimental study | **ChatGPT**, **Gemini,** **DeepSeek** | LLM-generated MI psychotherapy summaries | Highlighted both promise and risks of LLM summarization, especially semantic drift |
| **Batkhina, 2025 (USA)**[11] | **Psychotherapy / session notes** | **70 licensed psychotherapists in US**, client follow-up over **1 month** | **Pilot randomized controlled trial** | **Yung Sidekick**; NLP pipeline using **transformer-based models similar to BERT** trained on de- identified therapy transcripts | Automated therapy session note-taking by extracting **symptoms, goals, interventions, and medication mentions** from therapist input after sessions | Reduced time spent on session notes and preparation, improvements in adherence to treatment plans and perceived progress |
| **McCrudden et al., 2026 (USA)**[12] | **Mental health practice documentation / session notes** | Deidentified routine-care Talkspace session data; over **286,000 notes** generated across one year | Retrospective observational mixed-method study | **Smart Notes**; **Azure/OpenAI** with redaction pipeline (Amazon Glue/Python “scrubadub”) | Generative AI session-note drafting | Demonstrated large-scale real- world uptake of AI documentation support |
| **Ward et al., 2026 (USA)**[13] | **Psychotherapy / session notes** | Nationwide digital mental health benefit dataset: **8,724 providers**, **189,530 patients**, EHR- linked note & PHQ-9/GAD-7 outcome data | Retrospective cohort / large- scale observational evaluation | **Spring Health** teletherapy platform; session recording, **Amazon Transcribe**, LLM-based transcript summarization with prompting, de-identification pipeline | Therapy notetaking & summaries, especially the **“Presenting Problem”** and **“Session Focus”** sections | Demonstrated large-scale feasibility and adoption; AI- notetaking has **25% faster** note completion, **8.2 hours earlier** submission, and no adverse effect on continuity of care or depression/anxiety outcomes |

1. ASR: Automatic Speech Recognition; VADER: Valence Aware Dictionary and sEntiment Reasoner; NLP: Natural Language Processing;

2. BART: Bidirectional and Auto-Regressive Transformer; T5: Text-To-Text-Transformer

3. TaU: Treatment as Usual;

4. EMDR: Eye Movement Desensitization and Reprocessing;

5. MentalCLOUDS: Mental Health Counseling-Component–Guided Dialogue Summaries;

6. PIECE: Planning engine for mental counseling note generation; ACI-BENCH: a Novel Ambient Clinical Intelligence Dataset for Benchmarking Automatic Visit Note Generation;

7. MIMIC-III: Medical Information Mart for Intensive Care III; DENSE: Documenting Evolving Progress Notes from Scattered Evidence;

8. MI: Motivational Interviewing; MITI: Motivational Interviewing Treatment Integrity;
